# Statistical learning shapes pain perception and prediction independently of external cues

**DOI:** 10.1101/2023.03.23.23287656

**Authors:** Jakub Onysk, Nicholas Gregory, Mia Whitefield, Maeghal Jain, Georgia Turner, Ben Seymour, Flavia Mancini

**Affiliations:** Computational and Biological Learning Unit, Department of Engineering, University of Cambridge, Cambridge CB2 1PZ, UK; Applied Computational Psychiatry Lab, Max Planck Centre for Computational Psychiatry and Ageing Research, Queen Square Institute of Neurology and Mental Health Neuroscience Department, Division of Psychiatry, University College London, UK; MRC Cognition and Brain Sciences Unit, University of Cambridge, UK; Wellcome Centre for Integrative Neuroimaging, John Radcliffe Hospital, Headington, Oxford OX3 9DU, UK; Center for Information and Neural Networks (CiNet), Osaka 565-0871, Japan

## Abstract

The placebo and nocebo effects highlight the importance of expectations in modulating pain perception, but in everyday life we don’t need an external source of information to form expectations about pain. The brain can learn to predict pain in a more fundamental way, simply by experiencing fluctuating, non-random streams of noxious inputs, and extracting their temporal regularities. This process is called statistical learning. Here we address a key open question: does statistical learning modulate pain perception? We asked 27 participants to both rate and predict pain intensity levels in sequences of fluctuating heat pain. Using a computational approach, we show that probabilistic expectations and confidence were used to weight pain perception and prediction. As such, this study goes beyond well-established conditioning paradigms associating non-pain cues with pain outcomes, and shows that statistical learning itself shapes pain experience. This finding opens a new path of research into the brain mechanisms of pain regulation, with relevance to chronic pain where it may be dysfunctional.

## 1 INTRODUCTION

Clinical pain typically varies over time; in most pain states, the brain receives a stream of volatile and noisy noxious signals, which are also auto-correlated in time. The temporal structure of these signals is important, because the human brain has evolved the exceptional ability to extract regularities from streams of auto-correlated sensory signals, a process called statistical learning [1–7]. In the context of pain, statistical learning can allow the brain to predict future pain, which is crucial for orienting behaviour and maximising well-being [8, 9]. Statistical learning might also be fundamental to the ability of the nervous system to endogenously regulate pain. Indeed, statistical learning generates predictions about forthcoming pain. We already know that pain expectations can modulate pain levels by gating the reciprocal transmission of neural signals between the brain and spinal cord, as shown by previous work on placebo and nocebo effects [10–14].

By using temporal sequences of noxious inputs, we have previously shown that the pain system supports the statistical learning of the basic rate of getting pain by engaging both somatosensory and supramodal cortical regions [8]. Specifically, both sensorimotor cortical regions and the ventral striatum encode probabilistic predictions about pain intensity, which are updated as a function of learning by engaging parietal and prefrontal regions. According to a Bayesian inference framework, both the predictive inference and its confidence should, in principle, modulate the neural response to noxious inputs and affect perception, as a function of learning. In support of this conjecture, there is evidence that the confidence of probabilistic pain predictions modulates the cortical response to pain [9]. The relationship is inverse: the lower the confidence, the higher is the early cortical response to noxious inputs (and viceversa), as measured by EEG. This is expected based on Bayesian inference theory: when confidence is low, the brain relies less on his prior beliefs and more on sensory evidence to respond to the input. Bayesian inference theory also predicts that prior expectations and their confidence scale perception [15]. Thus, we hypothesise that the predictions generated by learning the statistics of noxious inputs in dynamically evolving sequences of stimuli modulate the perception of forthcoming inputs.

Previously, it was found that pain perception is strongly influenced by probabilistic expectations as defined by a cue that predicts high or low pain [16]. In contrast to such cue-paradigm, the primary aim of our experiment was to determine whether the expectations participants hold about the sequence itself inform their perceptual beliefs about the intensity of the stimuli. To that end, we recruited 27 healthy participants to complete a psycho-physical experiment where we delivered four different, 80-trial-long sequences of evolving thermal stimuli, with four levels of temporal regularity. On each trial, a 2s thermal stimulus was applied, following which participants were asked to either rate their perception of the intensity (Figure 1A) or to predict the intensity of the next stimulus in the sequence (Figure 1B). Participants also reported their response confidence.

**Figure 1.**
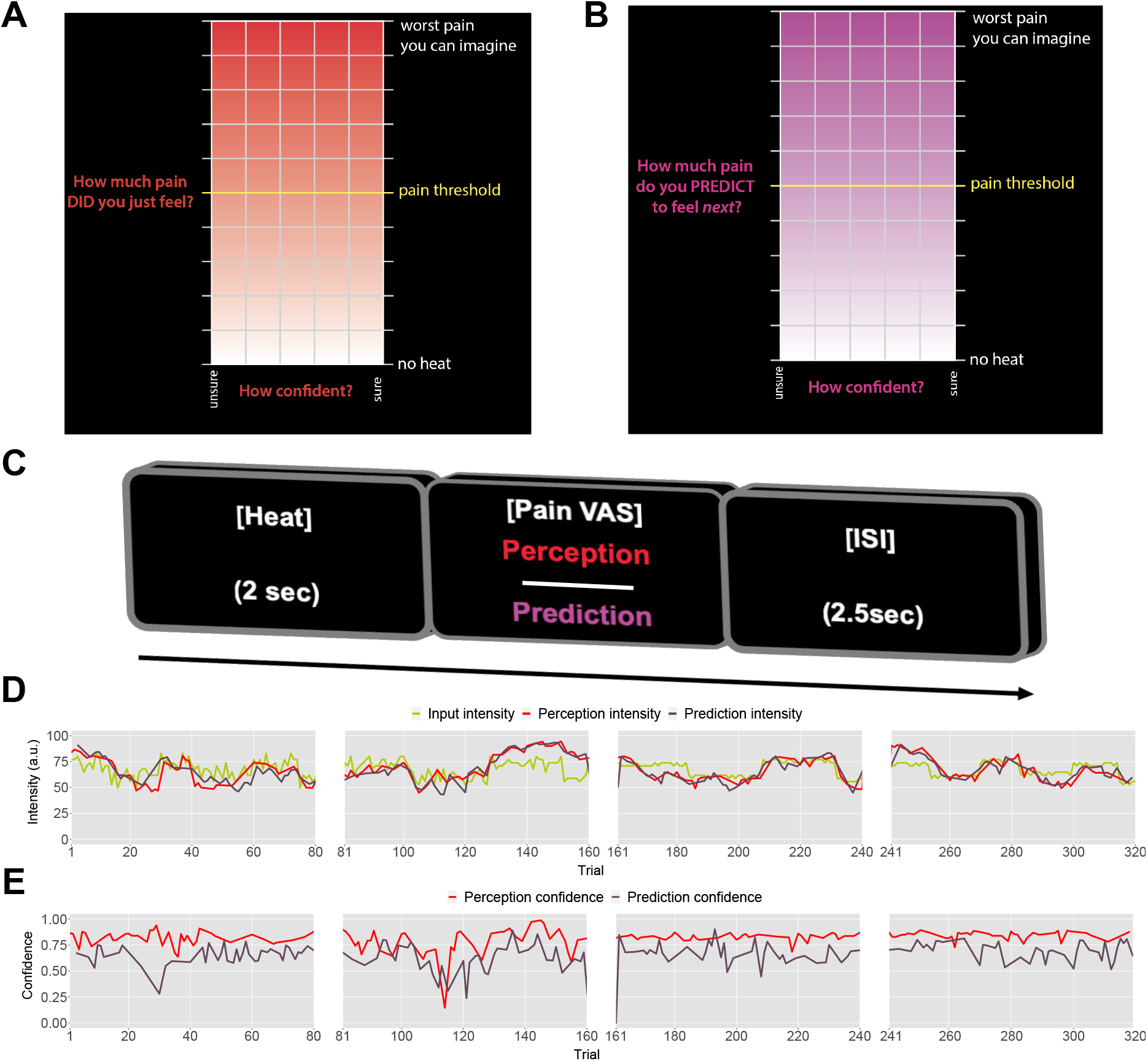
Task design. On each trial, each participant received a thermal stimulus lasting 2s from a sequence of intensities. This was followed by a perception (**A**) or a prediction (**B**) input screen, where the y-axis indicates the level of perceived/predicted intensity (0-100) centred around participant’s pain threshold, and the x-axis indicates the level of confidence in one’s perception (0-1). The inter-stimulus interval (ISI; black screen) lasted 2.5s (trial example in **C**). **D**: Example intensity sequences are plotted in green, participant’s perception and prediction responses are in red and black. **E**: Participant’s confidence rating for perception (red) and prediction (black) trials.

We contrasted four models of statistical learning, which varied according to the inference strategy used (i.e., optimal Bayesian inference or a heuristic) and the role of expectations on perception. All models used confidence ratings to weight the inference. We anticipate that probabilistic learning weighted by confidence and expectations modulates pain perception. This provides behavioural evidence for a link between learning and endogenous pain regulation. One reason why this is important is that it might help understand individual differences in the ability to endogenously regulate pain. This is particularly relevant for chronic pain, given that endogenous pain regulation can be dysfunctional in several chronic pain conditions [17–21], even before chronic pain develops [22]. Although there is ample evidence for changes in the functional anatomy and connectivity of endogenous pain modulatory systems in chronic pain, their computational mechanisms are poorly understood.

## 2 RESULTS

### 2.1 Model-naive performance

Prior to modelling, we first checked whether participant’s performance in the task was affected by the level of temporal regularity, i.e. the sequence condition. We varied the level of volatility and stochasticity across blocks (i.e. conditions), whilst we fixed their overall level within each block; the level of volatility was defined by the number of trials until the mean intensity level changes. The stochasticity is the additional noise that is added on each trial to the underlying mean, often referred to as the observation noise. The changes were often subtle and participants were not informed when they happened. We set two levels (low/high) of each type of uncertainty, achieving a 2x2 factorial design, with the order of conditions randomised across participants. A set of four example sequences of thermal intensities delivered to one of the participant’s can be found in Figure 1C, alongside their ratings of perception and predictions. Additionally, example confidence ratings for each type of response are plotted in Figure 1D. Figures S2-3 show the plots of each participant’s responses superimposed onto the sequences of noxious inputs.

As a measure of performance, we calculated the root mean squared error (RMSE) of participants responses (ratings and predictions) compared to the normative noxious input for each condition as in Figure 2 (see also Methods). The lower the RMSE, the more accurate participants’ responses are. Performance in different conditions was analysed with a repeated measures ANOVA, whose results are reported in full in Table S1. Although volatility did not affect rating accuracy (*F*(1, 26) = 0.96, *p* = 0.336, 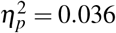), we found a 2-way interaction between the level of stochasticity of the sequence (low, high) and the type of rating provided (perceived intensity vs. prediction) (*F*(1, 26) = 29.842, *p <* 0.001, 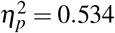). We followed up this interaction in post-hoc comparisons, as reported in Table S2. The performance score differences between all the pairs of stochasticity and response type interactions were significant, apart from the perception ratings in the stochastic environment as compared with perception and prediction performance in the low stochastic setting. Intuitively, the RMSE score analysis revealed an overall trend of participants performing worse on the prediction task, in particular when the

**Figure 2.**
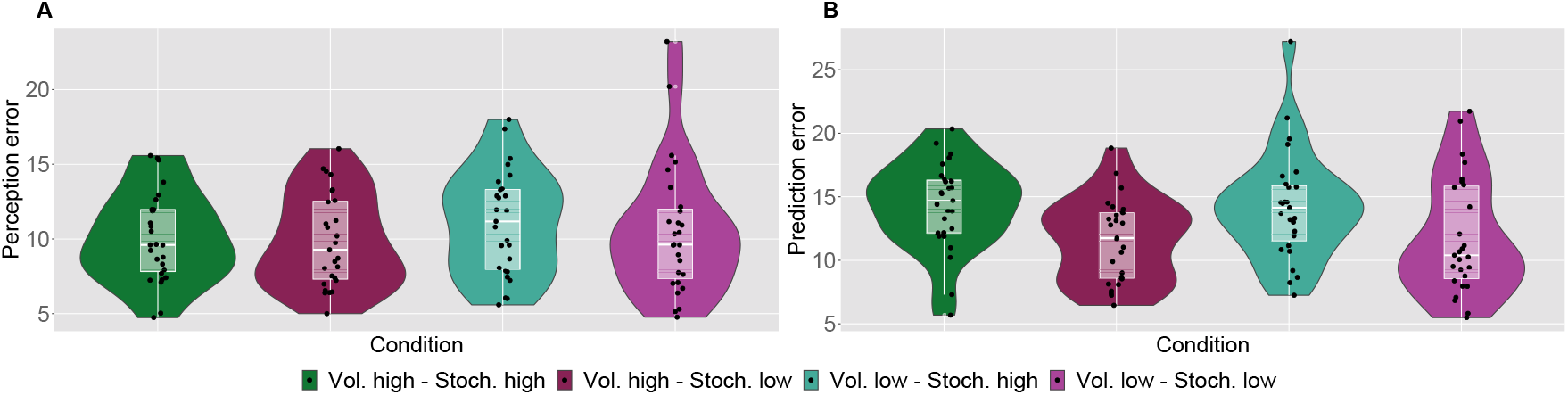
Participant’s model-naive performance in the task. Violin plots of participant Root Mean Square Error (RMSE) for each condition for **A**: rating and **B**: prediction responses as compared with the input.

### 2.2 Modelling strategy

Our models were selected a-priori, following the modelling strategy from [23] and hence considered the same set of core models for clear extension of the analysis to our non-cue paradigm. The key question for us was whether expectations were used to weight the behavioural estimates during sequence learning. Therefore, we compared Bayesian and non-Bayesian models of sequential learning that weighted their ratings based on prior expectations versus two corresponding models that assumed perfect perception (i.e. not weighted by prior beliefs). As a baseline, we included a random-response model (please see Methods for a formal treatment of the computational models).

According to an optimal Bayesian inference strategy, on each trial, participants update their beliefs about the feature of interest (thermal stimuli) based on probabilistic inference, maintaining a full posterior distribution over its values [16, 24]. Operating within a Bayesian paradigm, participants are assumed to track and, following new information, update both the mean of the sequence of interest and the uncertainty around it [25]. In most cases, such inference makes an assumption about environmental dynamics. For example, a common assumption is that the underlying mean (a hidden/latent state) evolves linearly according to a Gaussian random walk, with the rate of this evolution defined by the the variance of this Gaussian walk (volatility). The observed value is then drawn from another Gaussian with that mean, which has some observation noise (stochasticity). In this case, the observer can infer the latent states through the process of Bayesian filtering [24], using the Kalman Filter (KF) algorithm [26] (Figure 3B).

**Figure 3.**
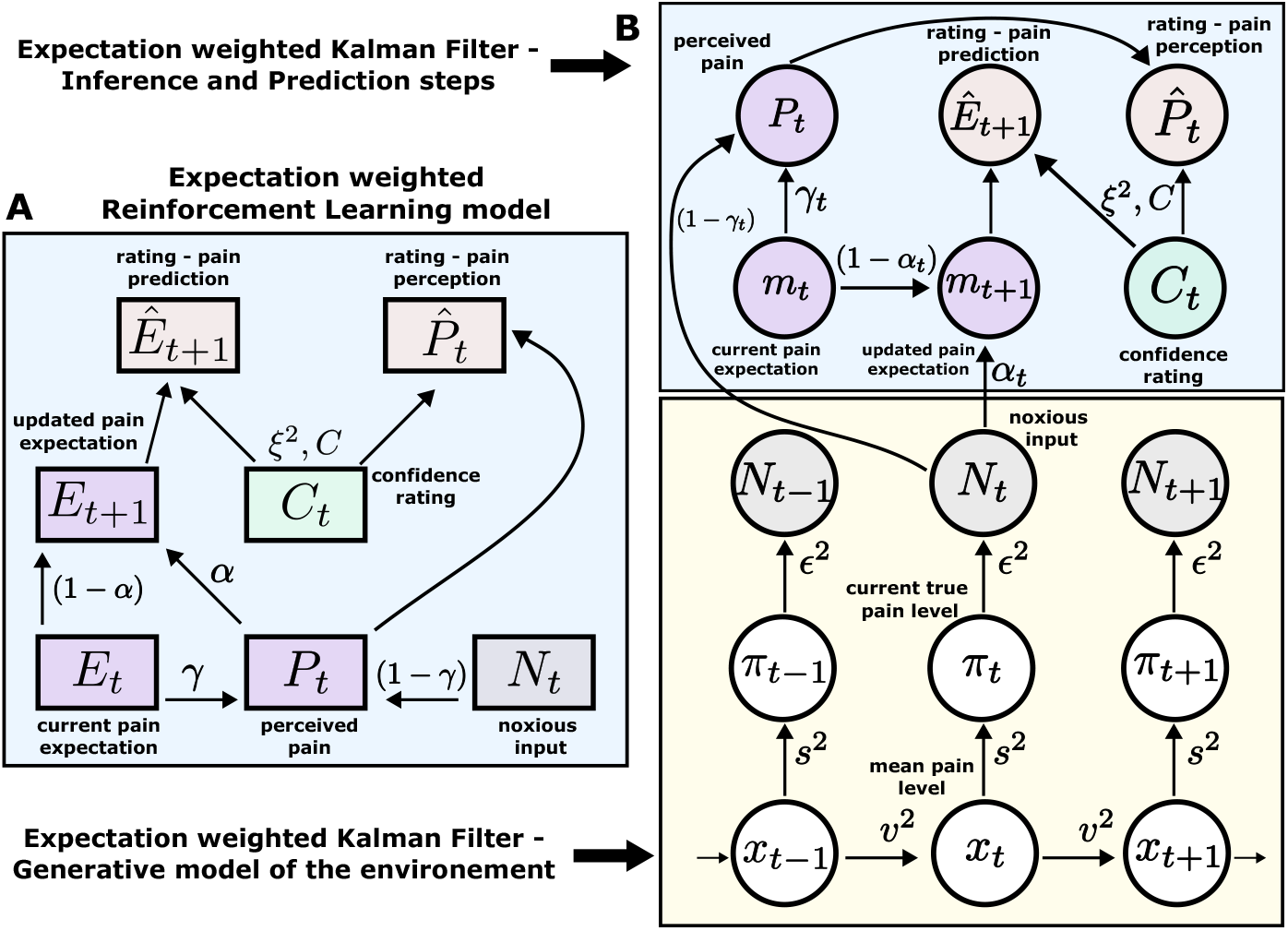
Expectation weighted models. Computational models used in the main analysis to capture participants’ pain perception 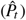 and prediction 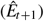 ratings. Both types of ratings are affected by confidence rating (*C*_*t*_) on each trial. **A**) In the Reinforcement Learning model, participant’s pain perception (*P*_*t*_) is taken to be weighted sum of the current noxious input (*N*_*t*_) and their current pain expectation (*E*_*t*_). Following the noxious input, participant updates their pain expectation (*E*_*t*+1_). (**B**) In the Kalman Filter model, a generative model of the environment is assumed (yellow background) - where the mean pain level (*x*_*t*_) evolves according to a Gaussian random walk (volatility *v*^2^). The true pain level on each trial (*π*_*t*_) is then drawn from a Gaussian (stochasticity *s*^2^). Lastly, the noxious input, *N*_*t*_, is assumed an imperfect indicator of the true pain level (subjective noise *ε*^2^). Inference and prediction steps are depicted in a blue box. Participant’s perceived pain is a weighted sum of expectation about the pain level (*m*_*t*_) and current noxious input (*N*_*t*_). Following each observation, *N*_*t*_, participant updates their expectation about the pain level (*m*_*t*+1_).

Sequence learning can also be captured by a heuristic to the Bayesian inference, i.e. a simple reinforcement learning (RL) rule. Here, participants maintain and update a point-estimate of the expected value of the sequence in an adaptive manner, within a non-stationary environment. RL explicitly involves correcting the tracked mean of the sequence proportionally to a trial-by-trial prediction error (PE) - a difference between the expected and actual value of the sequence [27] (Figure 3A). Importantly, RL agents do not assume any specific dynamics of the environment and hence are considered model-free.

Both models perform a form of error correction about the underlying sequence. The rate at which this occurs is captured by the learning rate *α* ∈ [0, 1] element. The higher the learning rate, the faster participants update their beliefs about the sequence after each observation. For the RL model, the learning rate *α* is a free parameter that is constant across the trials. On the other hand, the learning rate in the KF model *α*_*t*_ (known as the Kalman gain) is calculated on every trial. It depends on participants’ trial-wise belief uncertainty as well as their overall estimation of the inherent noise in the environment (stochasticity, *s*). The belief uncertainty is updated after each observation and depends on participants sense of volatility (*v*) and stochasticity (*s*) in the environment.

Crucially, we also used participants’ trial-by-trial confidence ratings to measure to what extent confidence plays a role in learning. This is captured by the confidence scaling factor *C*, which defines the extent to which confidence affects response (un-)certainty. Intuitively, the higher the confidence scaling factor *C*, the less important role confidence plays in participant’s response. With relatively low values of *C*, when the confidence is low, participants responses are more noisy, i.e. less certain. We demonstrate this in Figure 4 by plotting hypothetical responses (A-F) and the effect on the noise scaling (G-L) as a function of *C* and confidence ratings.

**Figure 4.**
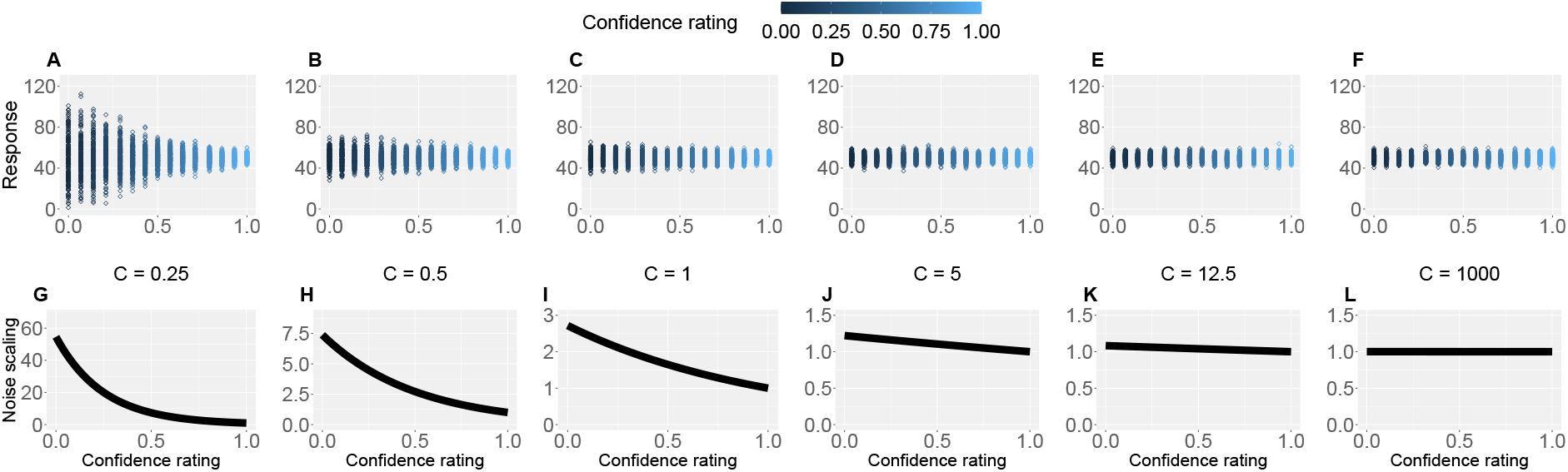
Confidence scaling factor demonstration. **A-F**: For a range of values of the confidence scaling factor *C*, we simulated a set of typical responses a participant would make for various levels of confidence ratings. The belief about the mean of the sequence is set at 50, while the response noise at 10. The confidence scaling factor *C* effectively scales the response noise, adding or reducing response uncertainty. **G-L**: The effect of different levels of parameter *C* on noise scaling. As *C* increases the effect of confidence is diminished.

To evaluate the effect of expectation on perceived intensity (on top of statistical learning modulating perception), we expanded the standard RL and KF models by adding a perceptual weighting element, *γ* ∈ [0, 1] (similarly to [16]). Essentially, *γ* governs how much each participant relies on the normative input on each trial, and how much their expectation of the input influences their reported perception - i.e. they take a weighted average of the two. The higher the *γ*, the bigger the impact of the expectation on perception. Again, in the case of the Reinforcement Learning model (eRL - expectation weighted RL), *γ* is a free parameter that is constant across trials, while in the Kalman Filter model (eKF - expectation weighted KF), *γ*_*t*_ is calculated on every trial and depends on: (1) the participants’ trial-wise belief uncertainty, (2) their overall estimation of the inherent noise in the environment (stochasticity, *s*) and (3) the participant’s subjective uncertainty about the level of intensity, *ε*.

Thus, in total we tested 5 models: RL and KF (perception not-weighted by expectations), eRL and eKF (perception weighted by expectations), and a baseline random model. We then proceeded to fit these 5 computational models to participants’ responses. For parameter estimation, we used hierarchical Bayesian methods, where we obtained group- and individual-level estimates for each model parameter (see Methods).

### 2.3 Modelling results

We fit each model for each condition sequence. Example trial-by-trial model prediction plots from one participant can be found in Figure S4. To establish which of the models fitted the data best, we ran model comparison analysis based on the difference in expected log point-wise predictive density (ELPD) between models. The models are ranked according to the ELPD (with the largest providing the best fit). The ratio between the ELPD difference and the standard error around it provides a significance test proxy through the sigma effect. We considered at least a 2 sigma effect as indication of a significant difference. In each condition, the expectation weighted models (eKF and eRL) provided better fit than models without this element (KF and RL; approx. 2-4 sigma difference, as reported in Figure 5A-D) and Table S5. This suggests that regardless of the levels of volatility and stochasticity, participants still weigh perception of the stimuli with their expectation. In particular, we found that the expectation-weighted KF model offered a better fit than the eRL, although in conditions of high stochasticity this difference was short of significance against the eRL model. This suggests that in learning about temporal regularities in the sequences of thermal stimuli, participants’ expectations modulate the perception of the stimulus. Moreover, this process was best captured by a model that updates the observer’s belief about the mean and the uncertainty of the sequence in a Bayesian manner.

**Figure 5.**
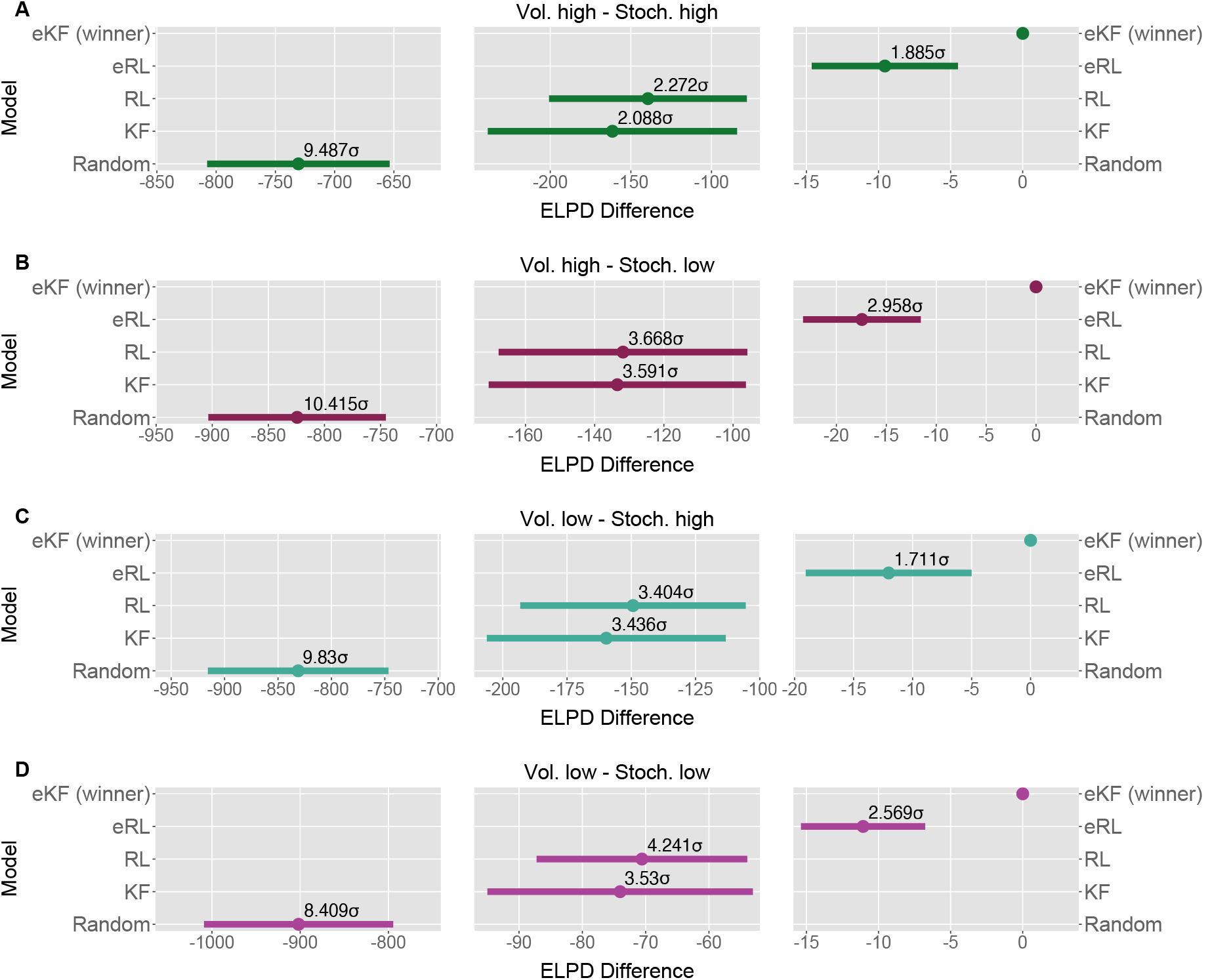
Model comparison for each sequence condition (**A-D**). The dots indicate the ELPD difference between the winning model (eKF) every other model. The line indicates the standard error (SE) of the difference. The non-winning models’ ELPD differences are annotated with the ratio between the ELPD difference and SE indicating the sigma effect, a significance heuristic.

We also found that as the confidence in the response decreases, the response uncertainty is scaled linearly with a negative slope ranging between 0.112-0.276 across conditions (Figure 6), confirming the intuition that less confidence leads to bigger uncertainty.

**Figure 6.**
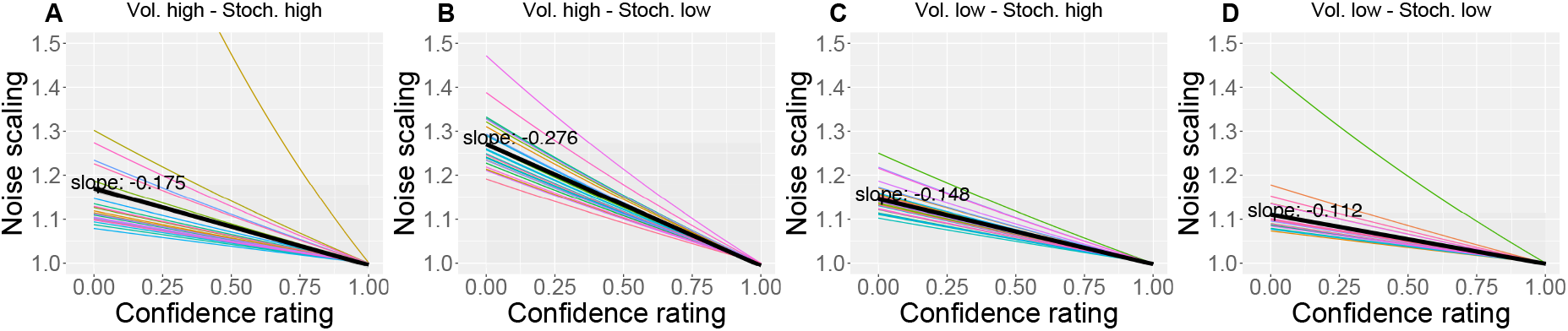
(**A-D**): The effect of the confidence scaling factor on noise scaling for each condition. Each coloured line corresponds to one participant, with the black line indicating the mean across all participants. The mean slope for each condition is annotated.

As an additional check, for each participant, condition and response type (perception and prediction), we plotted participants’ ratings against model predicted ratings and calculated a grand mean correlation in Figure S5. Next, we checked whether the parameters of the the winning eKF model differed across different sequence conditions. Given that volatility was fixed within condition, we treated it as a single-context scenario from the point of view of modelling [28], and we did not interpret its effect on the learning rate [29]. There were no differences for the group-level parameters; i.e., we did not detect significant differences between conditions in a hypothetical healthy participant group as generalised from our population of participants (Figure S12).

However, we found some differences at the individual-level of parameters (i.e. within our specific population of recruited participants), which we detected by performing repeated-measures ANOVAs (see Figure S13 for visualisation). The stochasticity parameter *s* was affected by the interaction between the levels of stochasticity and volatility (*F*(1, 26) = 35.108, *p <* 0.001, 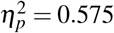), and was higher in highly stochastic and volatile conditions as compared to conditions where either volatility (*t* = 7.735, *p*_*bon f*_ *<* 0.001), stochasticity (*t* = 9.396, *p*_*bon f*_ *<* 0.001) or both were low (*t* = 8.826, *p*_*bon f*_ *<* 0.001). This suggests that, while participants’ performance was generally worse in highly stochastic environments, participants seem to have attributed this to only one source - stochasticity (*s*), regardless of the source of higher uncertainty in the sequence (stochasticity or volatility).

The response noise *ξ* was modulated by the level of volatility (*F*(1, 26) = 5.079, *p* = 0.033, 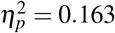), where it was smaller in highly volatile conditions. Moreover, we detected a significant interaction between volatility and stochasticity on the confidence scaling factor *C* (*F*(1, 26) = 81.258, *p <* 0.001, 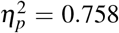), where the values *C* were overall lower when either volatility (*t* = −11.570, *p*_*bon f*_ *<* 0.001), stochasticity (*t* = −6.165, *p*_*bon f*_ *<* 0.001) or both (*t* = − 4.575, *p*_*bon f*_ *<* 0.001) were high as compared to the conditions where both levels of noise were low. This indicates there may have been some trade-off between *ξ* and *C*, as lower values of *C* introduce additional uncertainty when participant’s confidence is low.

Lastly, we found the initial uncertainty belief *w*_0_ was affected by the interaction between volatility and stochasticity (*F*(1, 26) = 5275.367, *p <* 0.001, 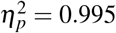) without a consistent pattern. All the other effects were not significant.

In summary, we formalised the process behind pain perception and prediction in noxious time-series within the framework of sequential learning, where the best description of participants’ statistical learning was captured through Bayesian filtering, in particular using a confidence-weighted Kalman Filter model. Most importantly, we discovered that, in addition to weighing their responses with confidence, participants used their expectations about stimulus intensity levels to form a judgement as to what they perceived. This mechanism was present across various levels of uncertainty that defined the sequences (volatility and stochasticity).

## 3 DISCUSSION

Statistical learning allows the brain to extract regularities from streams of sensory inputs and is central to perception and cognitive function. Despite its fundamental role, it has often been overlooked in the field of pain research. Yet, chronic pain appears to fluctuate over time. For instance, [30–32] reported that chronic back pain ratings vary periodically, over several seconds-minutes and in absence of movements. This temporal aspect of pain is important because periodic temporal structures are easy to learn for the brain [1, 8]. If the temporal evolution of pain is learned, it can be used by the brain to regulate its responses to forthcoming pain, effectively shaping how much pain it experiences. Indeed, we show that healthy participants extract temporal regularities from sequences of noxious stimuli and use this probabilistic knowledge to form confidence-weighted judgements and predictions about the level of pain intensity they experience in the sequence. We formalised our results within a Bayesian inference framework, where the belief about the level of pain intensity is updated on each trial according to the the amount of uncertainty participants ascribe to the stimuli and the environment. Importantly, their perception and prediction of pain were influenced by the expected level of intensity that participants held about the sequence before responding. When varying different levels of inherent uncertainty in the sequences of stimuli (stochasticity and volatility), the expectation and confidence weighted models fitted the data better than models weighted for confidence but not for expectations (Figure 5A-D). The expectation-weighted bayesian (KF) model offered a better fit than the expectation-weighted, model-free RL model, although in conditions of high stochasticity this difference was short of significance. Overall, this suggests that participants’ expectations play a significant role in the perception of sequences of noxious stimuli.

### 3.1 Statistical inference and learning in pain sequences

The first main contribution of our work is towards the understanding of the phenomenon of statistical learning in the context of pain. Statistical learning is an important function that the brain employs across the lifespan, with relevance to perception, cognition and learning [6]. The large majority of past research on statistical learning focused on visual and auditory perception [1, 3, 4, 33], with the nociceptive system receiving relatively little attention [34]. Recently, we showed that the human brain can learn to predict a sequence of two pain levels (low and high) in a manner consistent with optimal Bayesian inference, by engaging sensorimotor regions, parietal, premotor regions and dorsal striatum [8]. We also found that the confidence of these probabilistic inferences modulates the cortical response to pain, as expected by hierarchical Bayesian inference theory [9]. Here we tested sequences with a much larger range of stimulus intensities to elucidate the effect of statistical learning and expectations on pain perception. As predicted by hierarchical Bayesian inference theory, we find that the pain intensity judgements are scaled by both expectations and confidence.

Hence, our work highlights the inferential nature of the nociceptive system [16, 34–37], where in addition to the sheer input received by the nociceptors, there is a wealth of a priori knowledge and beliefs the agent holds about themselves and the environment that need to be integrated to form a judgement about pain [38–41]. This has an immediate significance for the real world, where weights need to be assigned to prior beliefs and/or stimuli to successfully protect the organism from further damage, but only to an extent to which it is beneficial.

Secondly, our results regarding the effect of expectation on pain perception relate to a much larger literature on this topic. The prime example would be placebo analgesia (i.e. the expectation of pain relief decreasing pain perception) and nocebo hyperalgesia (i.e. the expectation of high level of pain increasing its perception; [10, 37, 42, 43]. Recent work attempted to capture such expectancy effects within the Bayesian inference framework. For example, [25] showed that in addition to expectation influencing perceived pain in general, higher level of uncertainty around that expectation attenuated its effect on perception. Similarly, [44] demonstrated that when the discrepancy between the expectation and outcome (prediction error) is unusually large, the role of expectation is significantly reduced and so the placebo and nocebo effects are not that strong. An unusually large prediction error could be thought of as contributing to increased uncertainty about the stimuli, which mirrors the results from [25] Bayesian framework. Nevertheless, the types of stimuli used in the above studies (i.e. noxious stimuli cued by non-noxious stimuli) differed from the more ecologically valid sequences of pain that are reported by chronic pain patients [30], as we indicated above. Furthermore, [16] used a conditioning paradigm and also found that expectations influence both perception and learning, in a self-reinforcing loop. Our work has followed a similar modelling strategy to [16], but it goes beyond simple conditioning schedules or sequences of two-level discrete painful stimuli, showing expectancy effects even when the intensities are allowed to vary across a wider range of values and according to more complex statistical temporal structures. Additionally, given the reported role of confidence in the perception of pain [9, 45], we draw a more complete picture by including participants confidence ratings in our modelling analysis.

Future studies would need to determine whether statistical learning and its effect on pain is altered in chronic pain conditions. This is important because statistical learning could, in principle, influence how a pain state evolves. Once a pain state is initiated, how an individual learns and anticipates the fluctuating pain signals may contribute to determine how well it can be regulated by the nervous system, thus affecting the severity and recurrence of pain flares. This, in turn, would affect whether aversive associations with the instigating stimulus are extinguished or reinforced [36]. In chronic pain, dysfunctional learning may promote the amplification and maintenance of pain signals, contributing to the reinforcement of aversive associations with incident stimuli, as well as the persistence of pain [46–48].

Our paper comes with open tools, which can be adapted in future studies on statistical learning in chronic pain. The key advantage of taking an hypothesis-driven, computational-neuroscience approach to quantify learning is that it allows to go beyond symptoms-mapping, identifying the quantifiable computational principles that mediate the link between symptoms and neural function.

In summary, we show that statistical expectations and confidence scale the judgement of pain in sequences of noxious stimuli as predicted by hierarchical Bayesian inference theory, opening a new avenue of research on the role of learning in pain.

## 4 MATERIALS AND METHODS

### 4.1 Participants

Thirty-three (18 female) healthy adult participants were recruited for the experiment. The mean age of participants was 22.4 ± 2.7 years old (range: 18-35). Participants had no chronic condition and no infectious illnesses, as well as no skin conditions (e.g. eczema) at the site of stimulus delivery. Moreover, we only recruited participants that had not taken any anti-anxiety, anti-depressive medication, nor any illicit substances, alcohol and pain medication (including NSAIDs such as ibuprofen and paracetamol) in the 24 hours prior to the experiment. All participants gave informed written consent to take part in the study, which was approved by the local ethics committee.

### 4.2 Protocol

The experimental room’s temperature was maintained between 20°C to 23°C. Upon entry, an infrared thermometer was used to ensure participants temperature was above 36°C at the forehead and forearm of the non-dominant hand, to account for the known effects of temperature on pain perception [49]. A series of slideshows were presented, which explained the premise of the experiment and demonstrated what the participant would be asked to carry out. Throughout this presentation, questions were asked to ensure participants understood the task. Participants were given multiple opportunities to ask questions throughout the presentation.

We used the Medoc Advanced Thermosensory Stimulator 2 (TSA2) [50] to deliver thermal stimuli using the CHEPS thermode. The CHEPS thermode allowed for rapid cooling (40°C / sec) and heating (70°C / sec) so transitions between the baseline and stimuli temperatures were minimal. The TSA2 was controlled externally, via Matlab (Mathworks).

We then established the pain threshold, using the method of limits [51], in order to centre the range of temperature intensities used in the experiment. Each participant was provided with stimuli of increasing temperature, starting from 40°C going up in 0.5°C increments, using an inter-stimulus interval (ISI) of 2.5 sec and and a 2 sec duration. The participant was asked to indicate when the stimuli went from warm to painful - this temperature was noted and the stimuli ended. The procedure was repeated three times, and the average was used as an estimate of the pain threshold.

During the experiment, four sequences of thermal stimuli were delivered. Due to the phenomenon of offset analgesia (OA), where decreases in tonic pain result in a proportionally larger decrease in perceived pain [52], we chose phasic stimuli, with a duration of 2 sec and an inter-stimulus-interval (ISI) 2.5 seconds. In order to account for individual differences, the temperatures which the levels refer to are based upon the participants threshold.

The median intensity level was defined as threshold, giving a max temperature of 3°C above threshold, which was found to be acceptable by participants. Before the start of the experiment each participant was provided with the highest temperature stimuli that could be presented, given their measured threshold, to ensure they where comfortable with this. Two participants found the stimulus too painful - the temperature range was lowered by 1°C and this was found to be acceptable.

After every trial of each sequence, the participant was asked for either their perception of the previous stimulus, or their prediction for the next stimulus through a 2D VAS (Figure 1B), presented using PsychToolBox-3 [53]. The y-axis encodes the intensity of the stimulus either perceived or predicted, ranging from 0 (no heat detected/predicted) to 100 (worst pain imaginable perceived/predicted); on this scale, 50 represents the pain threshold. This was done as a given sequence was centred around the threshold. The x-axis encodes confidence in either perception or prediction, ranging from 0 - completely uncertain (‘unsure’) - to 1 - complete confidence in the rating (‘sure’). Differing background colours were chosen to ensure participants were aware of what was being asked, and throughout the experiment participants were reminded to take care in answering the right question. The mouse movement was limited to be inside of the coloured box, which defined the area of participants’ input. At the beginning of each input screen, the mouse location was uniformly randomised within the input box.

The sequence of response types was randomised so as to retain 40 prediction and 40 perception ratings for each of the four sequence conditions. For an 80-trial long sequence, this gave 80 participant responses. Each sequence condition was separated by a 5 minute break, during which the thermode’s probe was slightly moved around the area of skin on the forearm to reduce sensitisation (i.e. a gradual increase in perceived intensity with repetitive noxious stimuli) [54]. In the middle of each sequence, there was a 3 minute break. During the ISI, the temperature returned to a base-line of 38°C. One participant was unable to complete the sequence as their threshold was too low, and data from four participants was lost due to Medoc software issues (the remote control failed and the data of 2 out 4 sessions were not saved). We excluded one participant’s whose ratings/predictions were inversely proportional to the noxious input. Thus, we analysed data from 27 participants.

#### 4.2.1 Generative process of the painful sequences

We manipulated two sources of uncertainty in the sequence: the stochasticity (*s*) of the observation and the volatility (*v*) of the underlying sequence [29]. Sequences were defined by two levels (high or low) of stochasticity and volatility, resulting in four different sequences conditions - creating a 2x2 factorial design. Each sequence was defined as a series of chunks, where the intensity for trial *t, i*_*t*_ was sampled from *𝒩* (*I, σ* ^2^), where *σ* ^2^ indicates the level of stochasticity (*σ* ^2^ = 1.75 for high level of stochasticity, *σ* ^2^ = 0.25 for low level of stochasticity). The mean of the chunk, *I*, was drawn from *𝒰* (3.5, 10.5). To ensure a noticeable difference in chunk intensity to the participant, concurrent chunk means were constrained to be at least 2 intensity levels different. Volatility was implemented by defining the length, or number of trials, of a chunk (*l*) drawn from *𝒰* (*L − a, L* + *a*), where *L* is the mean o the chunk length (*L* = 15 for high volatility level, *L* = 25 for low volatility level). A jitter, *a*, was added around the mean to ensure the transition from one chunk to the next was not consistent or predictable. For both high and low volatility conditions we set *a* = 3. Sampled values were then discretised, where any intensities outside the valid intensity range [1, 13] were discarded and re-sampled resulting in an 80-trial long sequence for each condition. The mean of each sequence was centred around intensity level 7, i.e. the participants threshold. So defined, six sets of four sequences were sampled. Each participant received one set, with a randomised sequence order. See an example sequence (after subject-specific linear transformation) and one participant’s responses (including confidence ratings) in Figure 1C-D.

### 4.3 Data pre-processing

Since the intensity values of the noxious input were discretised between 1 and 13, while the participant’s responses (perception and prediction) were given on a 0-100 scale, we applied a linear transformation of the input to map its values onto a common 0-100 range. For each participant, for a set of inputs at perception trials from the concatenated sequence (separate sequence conditions in the order as presented), we fit a linear least-squares regression using Python’s `scipy.stats.linregress` function. On rare occasions, when the transformed input was negative, we refit the line using Python’s non-linear least squares function `scipy.optimize.curve_fit`, constraining the intercept above 0 [55]. We then extracted each participant’s optimised slope and intercept and applied the transformation both to the concatenated and condition-specific sequence of inputs. So transformed, the sequences where then used in all the analyses. Plots of each participant transformation can be found in Figure S1. We superimposed participant’s responses onto the noxious input condition sequences in Figure S2.

To capture participant’s model-naive performance in the task, both for the concatenated and condition-specific sequence, we calculated Root Mean Square Error (RMSE) of each participant’s perception (Eq. 1) and prediction (Eq. 2) responses as compared to the input. The lower the RMSE, the higher the response accuracy.

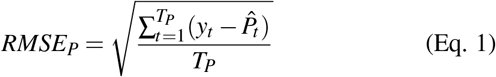

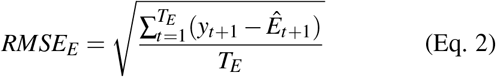

where *T*_*P*_ is the number of perception trials, 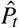 is participant’s perception response to the stimulus *y*_*t*_ at trial *t, T*_*E*_ is the number of prediction trials and 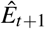 is participant’s prediction of the next stimulus intensity *y*_*t*+1_ at at trial *t* + 1.

### 4.4 Models

#### 4.4.1 Reinforcement Learning

##### RL

In reinforcement learning models, learning is driven by discrepancies between the estimate of the expected value and observed values. Before any learning begins, at trial *t* = 1, participants have an initial expectation, *E*_1_ = *E*_0_, which is a free parameter that we estimate.

On each trial, participants receive a thermal input *N*_*t*_. We then calculate the prediction error *δ*_*t*_, defined as the difference between the expectation *E*_*t*_ and the input *N*_*t*_ (Eq. 3).

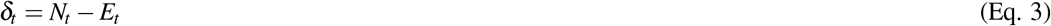

Participant is then assumed to update their expectation of the stimulus on the next trial as in Eq. 4

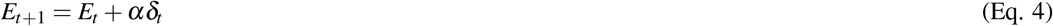

where *α* is the learning rate (free parameter), which governs how fast participants assimilate new information to update their belief.

On trials when participants rate their perceived intensity, we assume no effects on their perception other than confidence rating *c*_*t*_ and response noise, so participants perception response 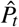 is drawn from a Gaussian distribution, with the mean *P*_*t*_ = *N*_*t*_ and a confidence-scaled response noise *ξ* (free parameter), as in Eq. 5

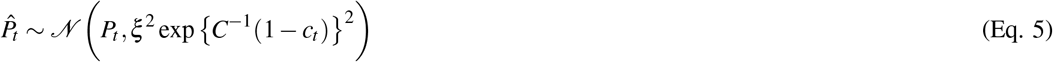

where *C* is the confidence scaling factor (free parameter), which defines the extent to which confidence affects response uncertainty. Please, see Figure 4 for an intuition behind confidence scaling.

On trials when participants are asked to predict the intensity of the next thermal stimulus, we use the updated expectation *E*_*t*+1_ to model participants prediction response 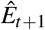. This is similarly affected by confidence rating and response noise and is defined as in Eq. 6.

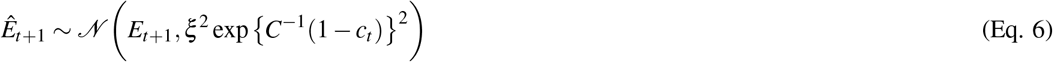

To recap, the RL model has 4 free parameters: the learning rate *α*, response noise *ξ*, the initial expectation *E*_0_, and the confidence scaling factor *C*.

##### eRL

Additionally, where we investigate the effects of expectation on the perception of pain [16], we included an element that allows us express the perception as a weighted sum of the input and expectation (Eq. 7)

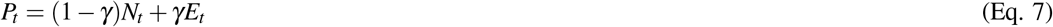

where *γ* ∈ [0, 1] (free parameter) captures how much participants rely on the normative thermal input vs. their expectation. When *γ* = 0, the expectation plays no role and the model simplifies to that of the standard RL above. In total, the eRL model has 5 free parameters, with the other equations the same as in the RL model, with the exception of the prediction error, which now relies on the expectation-weighted pain perception *P*_*t*_ (Eq. 8).

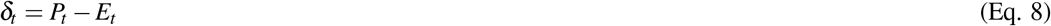

#### 4.4.2 Kalman Filter

##### KF

To capture sequential learning in a Bayesian manner, we used the Kalman filter model [16, 24, 26]. KF assumes a generative model of the environment where the latent state on trial t, *x*_*t*_ (the mean of the sequences in the experiment), evolves according to a Gaussian random walk with a fixed drift rate, *v* (volatility), as in Eq. 9.

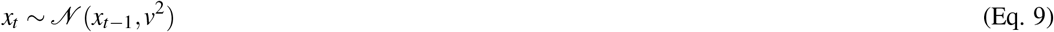

The observation on trial t, *N*_*t*_, is then drawn from a Gaussian (Eq. 10) with a fixed variance, which represents the observation uncertainty *s* (stochasticity).

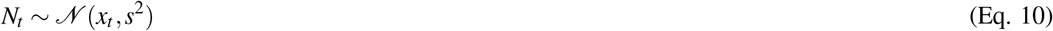

As such the KF assumes stable dynamics since the generative process has fixed volatility and stochasticity.

For ease of explanation, we refer to the thermal input at each trial as *N*_*t*_, we also use the *N*_1:*t*_ notation, which refers to a sequence of observations up to and including trial *t*. The model allows to obtain posterior beliefs about the latent state *x*_*t*_ given the observations. This is done by tracking an internal estimate of the mean *m*_*t*_ and the uncertainty, *w*_*t*_, of the latent state *x*_*t*_.

First, following standard KF results, on each trial, the participant is assumed to hold a prior belief (indicated with ^(−)^ superscript) about the latent state, *x*_*t*_ (Eq. 11).

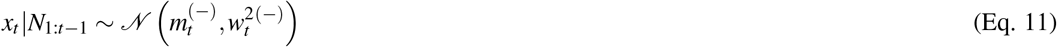

On the first trial, before any observations, we set 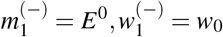 (free parameters). In light of the new observation, *N*_*t*_ on trial *t*, the tracked mean and uncertainty of the latent state are reweighed based on the new evidence *N*_*t*_ and its associated observation uncertainty *s* as in Eq. 12.

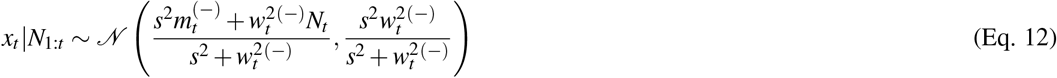

We can then define the learning rate *α*_*t*_ (Eq. 13),

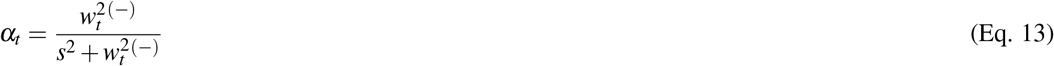

to get the update rule for the new posterior beliefs (indicated with ^(+)^ superscript) about the mean (Eq. 14) and uncertainty (Eq. 15) of *x*_*t*_.

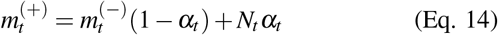

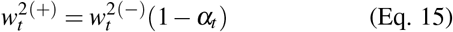

Following this new belief, and the assumption about the environmental dynamics (volatility), the participant forms a new prior belief about the latent state *x*_*t*+1_ for the next trial *t* + 1 as in Eq. 16.

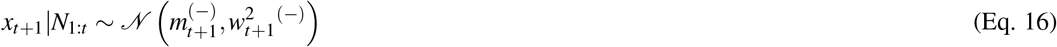

Where

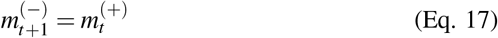

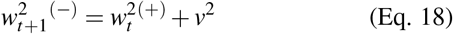

We can simplify the notation to make it comparable to the RL models. We let, 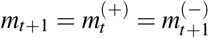, and 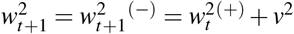. Following a new observation at trial *t*, we calculate the prediction error (Eq. 19) and learning rate (Eq. 20).

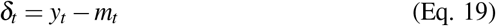

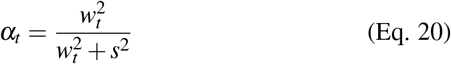

we then update the belief about the mean (Eq. 21) and uncertainty (Eq. 22) of the latent state for the next trial.

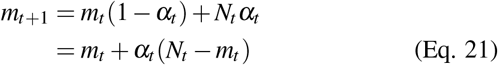

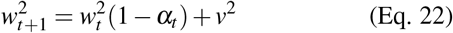

Now, mapping this onto the experiment, the mean of the latent state is participants expectation *E*_*t*_ = *m*_*t*_, and so we have participant perception rating modelled as in Eq. 23.

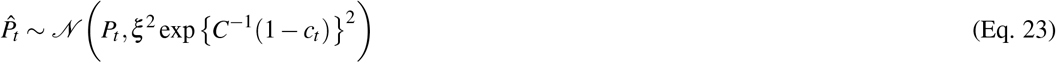

and the prediction rating for the next trial as in Eq. 24.

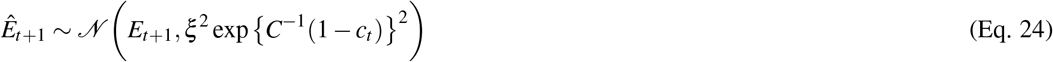

In total the model has 6 free parameters: *s* (environmental stochasticity), *v* (environmental volatility), *ξ* (response noise), *E*_0_ (initial belief about the mean), *w*_0_ (initial belief about the uncertainty) and *C* (confidence scaling factor).

##### eKF - expectation weighted Kalman Filter

We can introduce the effect of expectation on the pain perception, by assuming that participants treat the thermal input as an imperfect indicator of the true level of pain [16]. In this case, the input, *N*_*t*_, is modelled as in Eq. 25

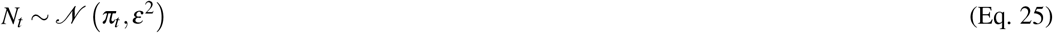

which forms an expression for the likelihood of the observation and adds an additional level to the inference, slightly modifying the Kalman filter assumptions such that:

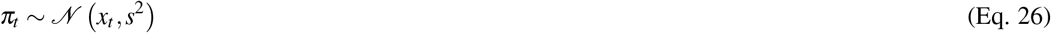

However, we can apply the standard KF results and Bayes’ rule to arrive at simple update rules for the participants’ belief about the mean and uncertainty of the latent state *x*_*t*_. From this, we get a prior on the *π*_*t*_ defined in Eq. 27

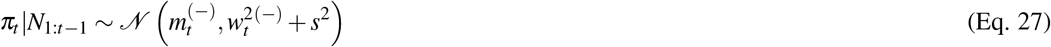

which, following a new input *N*_*t*_, gives us the posterior belief about *π*_*t*_ as in Eq. 28.

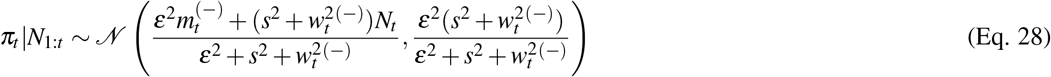

Now, if we define *γ*_*t*_ as in Eq. 29

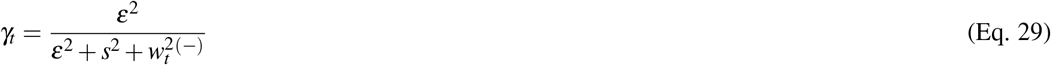

we have that the posterior belief about the mean level of pain *π*_*t*_ is calculated as:

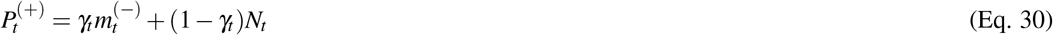

which is a weighted sum of the input *N*_*t*_ and participant expectation about the latent state *x*_*t*_, governed by the perceptual weight *γ*_*t*_, analogously to the eRL model. Finally, the posterior belief about *x*_*t*_ is obtained in Eq. 31.

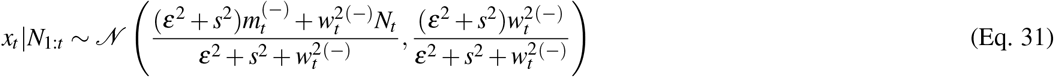

Now, setting the learning rate as in Eq. 32

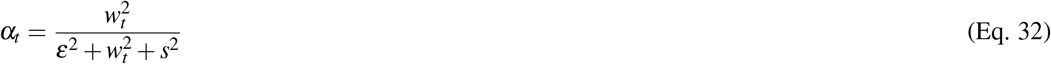

we get:

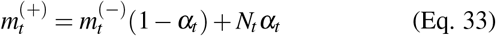

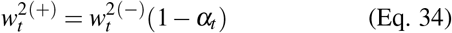

Next, following the same notation simplification as before, we get the update rules for the prior belief about the mean (Eq. 35) and uncertainty (Eq. 36) of the latent state *x*_*t*+1_ for the next trial.

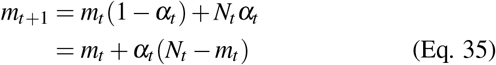

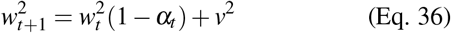

as well as the expression for subjective perception, *P*_*t*_, at trial *t* (Eq. 37).

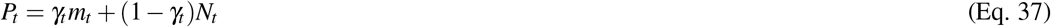

The perception and prediction responses are modelled analogously as the KF model. In total, the model has 7 free parameters: *ε* (subjective noise), *s* (environmental stochasticity), *v* (environmental volatility), *ξ* (response noise), *E*_0_ (initial belief about the mean), *w*_0_ (initial belief about the uncertainty) and *C* (confidence scaling factor).

#### 4.4.3 Random model

As a baseline, we also included a model that performs a random guess. The perceptual/prediction ratings were modelled as in Eq. 38.

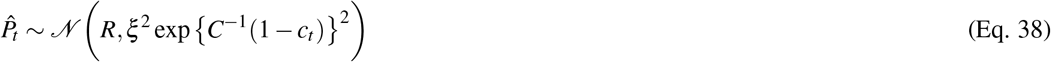

The model has 3 free parameters: *R, ξ*, and *C*, where *R* is a constant value that participants respond with.

### 4.5 Model fitting

Model parameters were estimated using hierarchical Bayesian methods, performed with RStan package (v. 2.21.0) [56] in R (v. 4.0.2) based on Markov Chain Monte Carlo techniques (No-U-Turn Hamiltonian Monte Carlo). For the individual level-parameters we used non-centred parametrisation [57]. For the group-level parameters we used *𝒩* (0, 1) priors for the mean, and the gamma-mixture representation of the Student-t(3,0,1) for the scale [58]. Parameters in the (0, 1) range were constrained using `Phi_approx` - a logistic approximation to the cumulative Normal distribution [59].

For each condition and each of the four chains, we ran 6000 samples (after discarding 6000 warm-up ones). For each condition, we examined R-hat values for each individual- (including the *𝒩* (0, 1) error term from the non-centred parametrisation) and group-level parameters from each model to verify whether the Markov chains have converged. At the group-level and individual-level, all R-hat values had a value *<* 1.1, indicating convergence. In the random response model, 0.01% − 0.16% iterations saturated the maximum tree depth of 11.

#### 4.5.1 Model comparison

For model comparison, we used R package `loo`, which provides efficient approximate leave-one-out cross-validation. The package allows to estimate the difference in models’ expected predictive accuracy through the difference in expected log point-wise predictive density (ELPD) [60]. By looking at the ratio between the ELPD difference and the standard error (SE) of the difference, we get the sigma effect - a heuristic for significance of such model differences. There’s no agreed-upon threshold of SEs that determines significance, but the higher the sigma difference, the more robust is the effect. The closeness of fit can be also captured with LOO information criterion (LOOIC), where the lower LOOIC values indicate better fit.

#### 4.5.2 Parameter comparison

For the comparison of group-level parameters between conditions,we extracted 95% High Density Intervals (HDI) of the permuted and merged (across chains) posterior samples of each group-level parameter [61]. To assess significant differences between conditions, we calculated a difference between such defined intervals. In the Bayesian scenario, a significant difference is indicated by the interval not containing the value 0 [62, 63].

#### 4.5.3 Parameter and model recovery

To asses the reliability of our modelling analysis [64], for each model we performed parameter recovery analysis, where we simulated participants’ responses using newly drawn individual-level parameters from the group-level distributions.

We repurposed existing sequences of noxious inputs in the [1, 13] range (pre-transformation). When then applied a linear transformation to the input sequences using sampled slope and intercept coefficients from a Gaussian distribution of these coefficients that we estimated based on our dataset using R’s `fitdistrplus` package. Furthermore, we simulated the confidence ratings based on lag-1 auto-correlation across a moving window of the transformed input sequence.

We then fit the same model to the simulated data and calculated Pearson correlation coefficients *r* between the generated and estimated individual-level parameters. The higher the coefficient *r*, the more reliable the estimates are, which can be categorised as: poor (if r <0.5); fair (if 0.5< r <0.75); good (0.75< r <0.9); excellent (if r >0.9) [65]. Results are reported in Table S3 and Figures S6-11.

We also performed model recovery analysis [64], where we first simulated responses using each model and then fit each model-specific dataset with each model. We then counted the number of times a model fit the simulated data best (according to the LOOIC rule), effectively creating an *M* × *M* confusion matrix, where *M* is the number of models. In the case where we have a diagonal matrix of ones, the models are perfectly recoverable and hence as reliable as possible. Results are reported in Table S4.

In Tables S6-S9 we report Bulk and Tail Effective Sample Size (ESS) for each condition, for each model and parameter.

## Data Availability

All code and data will be available open source, released upon acceptance of the paper in a peer-reviewed journal.

## 5 DATA AND CODE AVAILABILITY

All code and data will be available open source, released upon acceptance of the paper.

## 6 ACKOWLEDGEMENTS

The study was funded by a MRC Career Development Award to FM (MR/T010614/1) and a UKRI Advanced Pain Discovery Platform grant to both F.M. and B.S. (MR/W027593/1). B.S. was also funded by Wellcome (214251/Z/18/Z), Versus Arthritis (21537), and IITP (MSIT 2019-0-01371). This work has been performed using resources provided by the Cambridge Tier-2 system operated by the University of Cambridge Research Computing Service (www.hpc.cam.ac.uk) funded by EPSRC Tier-2 capital grant (EP/T022159/1). HPC access was additionally funded by an EPSRC research infrastructure grant to F.M.. We are grateful to Professor Máté Lengyel and Professor Deborah Talmi for helpful discussions about the study. For the purpose of open access, the author has applied a Creative Commons Attribution (CC BY) licence to any Author Accepted Manuscript version arising from this submission.

## 7 AUTHOR CONTRIBUTIONS

JO, NG, MW, GT, BS and FM designed the study. NG collected the data. JO, NG, MJ and FM analysed the data. JO, NG, MW, BS and FM wrote the article.

