## Supplementary information for "Statistical learning shapes pain perception and prediction independently of external cues"

#### 1 Behavioural results

##### 1.1 Model-naive performance

For each sequence condition (Volatility × Stochasticity), we calculated the root mean squared error (RMSE) of participants responses (Type: perception and prediction ratings) and compared to the normative noxious input, as a measure of performance in the task. We analysed the RMSEs with a repeated measures ANOVA, with the results reported in Table S1.

Given the significant interaction between stochasticty and response type, we further ran a post hoc comparison tests for this effect, as reported in Table S2.

Table S1: Within subjects effects from repeated measures ANOVA of participant RMSE scores with stochasticity, volatility and response type factors. SS - Sum of Squares, MS - Mean Square.

| Effect | SS | df | MS | F | p | $\eta^2$ | $\eta^2_p$ |
|---|---|---|---|---|---|---|---|
| Volatility | 10.714 | 1 | 10.714 | 0.960 | 0.336 | 0.007 | 0.036 |
| Residuals | 290.166 | 26 | 11.160 | | | | |
| Stochasticity | 113.964 | 1 | 113.964 | 19.939 | $<$ **0.001*** | 0.074 | 0.434 |
| Residuals | 148.603 | 26 | 5.715 | | | | |
| Type | 365.000 | 1 | 365.000 | 85.109 | $<$ **0.001*** | 0.237 | 0.766 |
| Residuals | 111.503 | 26 | 4.289 | | | | |
| Volatility × Stochasticity | 0.006 | 1 | 0.006 | $5.688e-4$ | 0.981 | $3.723e-6$ | $2.188e-5$ |
| Residuals | 261.912 | 26 | 10.074 | | | | |
| Volatility × Type | 7.313 | 1 | 7.313 | 3.196 | 0.085 | 0.005 | 0.109 |
| Residuals | 59.487 | 26 | 2.288 | | | | |
| Stochasticity × Type | 63.662 | 1 | 63.662 | 29.842 | $<$ **0.001*** | 0.041 | 0.534 |
| Residuals | 55.466 | 26 | 2.133 | | | | |
| Volatility × Stochasticity × Type | 1.356 | 1 | 1.356 | 0.704 | 0.409 | $8.807e-4$ | 0.026 |
| Residuals | 50.060 | 26 | 1.925 | | | | |

Table S2: Post Hoc Comparisons for the repeated measures ANOVA's interaction effect of Stochasticity × Type.

| | | | 95% CI for Mean Diff. | | | | |
|---|---|---|---|---|---|---|---|
| | | Mean Diff. | Lower | Upper | SE | t | $p_{bonf}$ |
| High, Perception | Low, Perception | 0.367 | −0.687 | 1.421 | 0.381 | 0.963 | 1.000 |
| | High, Prediction | −3.686 | −4.636 | −2.735 | 0.345 | −10.688 | $<$ **0.001*** |
| | Low, Prediction | −1.147 | −2.329 | 0.034 | 0.430 | −2.665 | 0.062 |
| Low, Perception | High, Prediction | −4.053 | −5.234 | −2.871 | 0.430 | −9.415 | $<$ **0.001*** |
| | Low, Prediction | −1.514 | −2.464 | −0.564 | 0.345 | −4.390 | $<$ **0.001*** |
| High, Prediction | Low, Prediction | 2.539 | 1.484 | 3.593 | 0.381 | 6.658 | $<$ **0.001*** |

### 2 Noxious inputs and responses

#### 2.1 Input transformation

We linearly transformed participants' responses to project them from the 1-13 range to 0-100 using a linear transformation we obtained from a regression of stimulus intensities onto pain ratings.

Plots of each participant's transformation can be found in Figure S1.

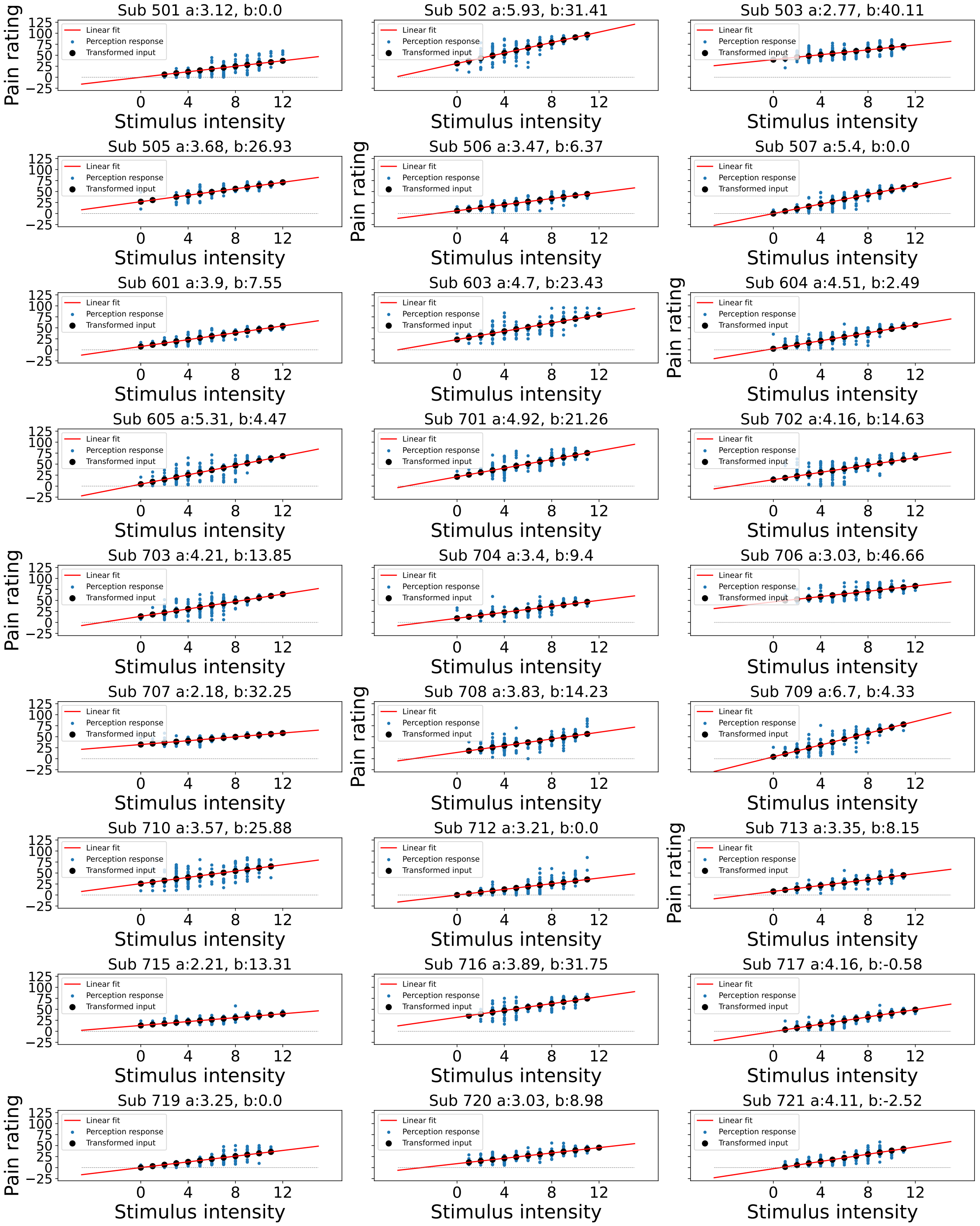


Figure S1: Linear transformation of the input at perception trials. Blue dots indicate participant's perception responses for a given level of stimulus intensity, black dots indicate transformed intensity values, a linear least squares regression was performed to achieve the best fitting line through participant responses as shown in red, the intercept was constrained >0.

We superimposed participants responses (perception and prediction ratings) onto the noxious input condition sequences in Figure S2. The black line marks the start of a new sequence condition.

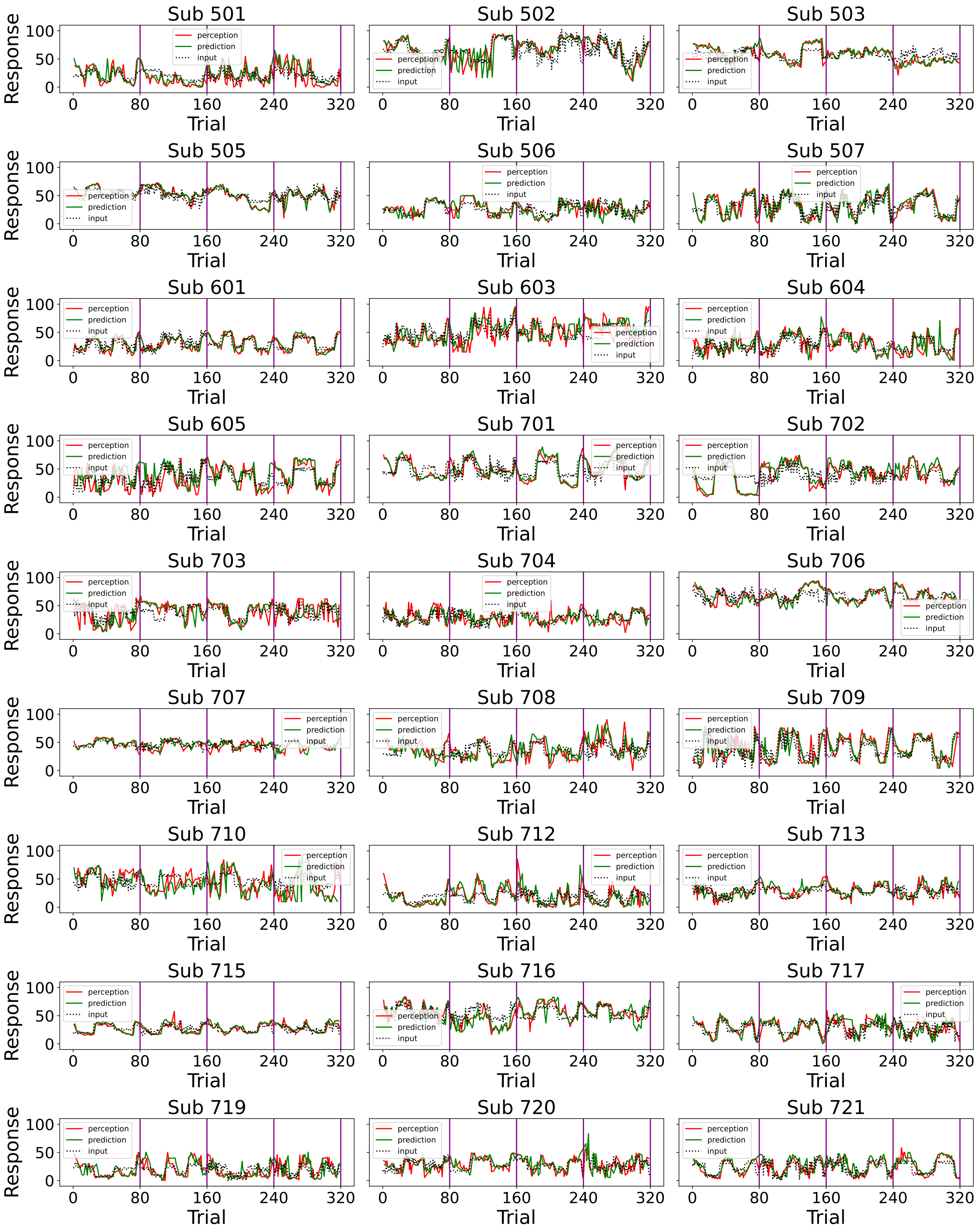


Figure S2: Participants responses (red - perception; green - prediction) to the noxious input (dotted line) sequences. Vertical purple lines mark the end of each condition.

Finally, we plotted participants' confidence ratings throughout the task in Figure S3.

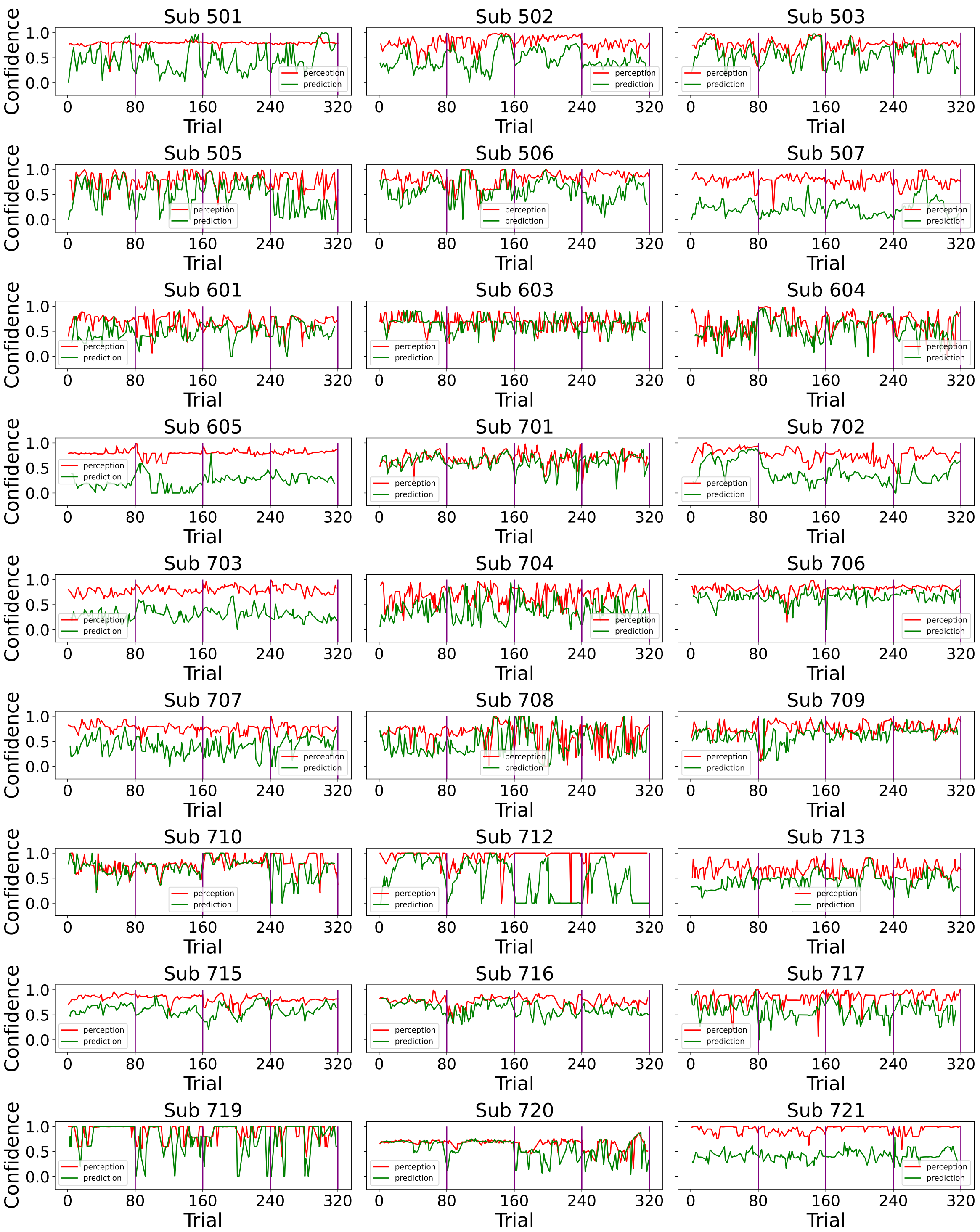


Figure S3: Participants' confidence ratings in (red - perception; green - prediction) during the task. Vertical purple lines mark the end of each condition.

### 3 Model predictions

Following the model fitting procedure, in Figure S4 we plotted example model predicted ratings both for perception and prediction responses for each condition as compared with noxious input for one participant's responses.

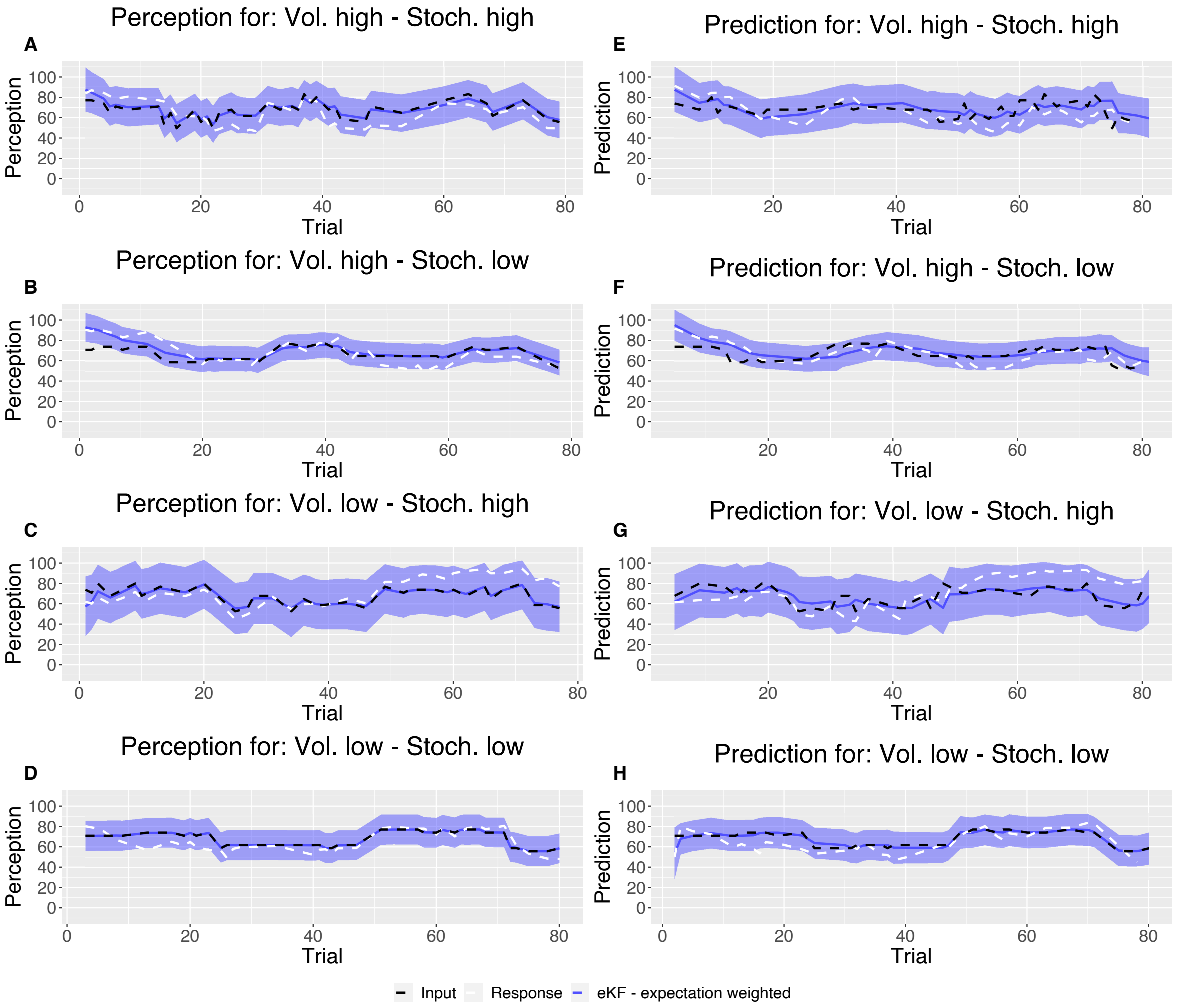


Figure S4: Example plot of the input sequences (black) for each condition, one participant's responses (white) and the winning, eKF, model predictions (blue) including 95% confidence intervals (shaded blue) for **A-D**: Perception and **E-H**: Prediction.

Lastly, for each condition and for each participant we plotted model responses (perception and prediction) against participant responses in Figure S5. The grand mean correlation across participants, for each condition and response type was calculated and included in the figure.

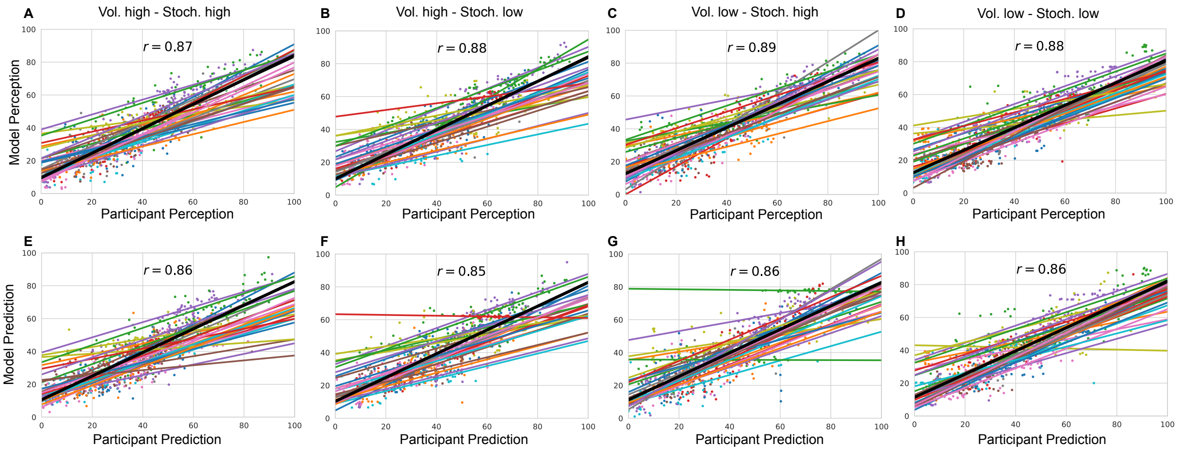


Figure S5: Model responses against participants responses for each condition and each response type **A-D**: Perception and **E-H**: Prediction. The annotated value is the grand mean correlation across subjects for each condition and response type.

### 4 Parameter recovery

The results of parameter recovery analysis for each parameter for each model are reported in Table S3. We recovered each individual (out 27 participants) parameter $\approx$ 100 times and calculated the mean and SD of the correlation between the true and recovered parameter values.

Table S3: Pearson correlation coefficient $r$ (SD) from the parameter recovery analysis for each model

| | | | | | | | |
|---|---|---|---|---|---|---|---|
| | eRL | | | | | | |
| | $\alpha$ | $\gamma$ | $\xi$ | $E^0$ | $C$ | | |
| $r$ (SD) | 0.685 (0.113) | 0.92 (0.049) | 0.993 (0.005) | 0.723 (0.093) | 0.481 (0.131) | | |
| | RL | | | | | | |
| | $\alpha$ | $\xi$ | $E^0$ | $C$ | | | |
| $r$ (SD) | 0.842 (0.081) | 0.993 (0.004) | 0.625 (0.107) | 0.455 (0.133) | | | |
| | eKF | | | | | | |
| | $\epsilon$ | $s$ | $v$ | $\xi$ | $E^0$ | $w^0$ | $C$ |
| $r$ (SD) | 0.742 (0.1) | 0.531 (0.13) | 0.745 (0.09) | 0.986 (0.075) | 0.849 (0.118) | 0.309 (0.179) | 0.472 (0.123) |
| | KF | | | | | | |
| | $s$ | $v$ | $\xi$ | $E^0$ | $w^0$ | $C$ | |
| $r$ (SD) | 0.605 (0.129) | 0.589 (0.117) | 0.993 (0.005) | 0.585 (0.157) | 0.298 (0.18) | 0.442 (0.146) | |
| | Random model | | | | | | |
| | $\xi$ | $R$ | $C$ | | | | |
| $r$ (SD) | 0.996 (0.004) | 0.999 (0.001) | 0.079 (0.206) | | | | |

Moreover, to assess the number of simulations needed, we calculated the average SD (and its error) of the correlation as a function of increasing number of simulation, as plotted in Figure S6. The average was obtained from the 500 randomly chosen permutations of different simulations at each $n$ (out of $\approx$ 100).

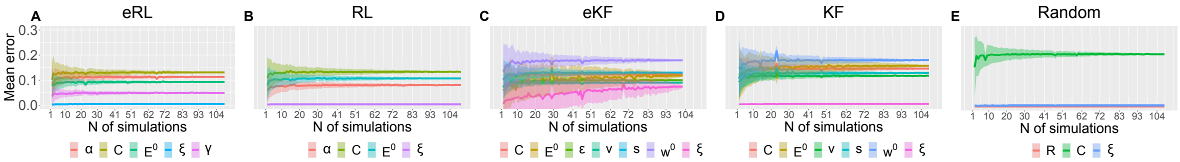


Figure S6: **Parameter recovery average SD for each model.** The average SD is plotted as a function of simulation number averaged across 500 permutations of ≈ 100 simulations.

Next, we include scatter plots from the parameter recovery for each model and parameter in Figures S7-S11.

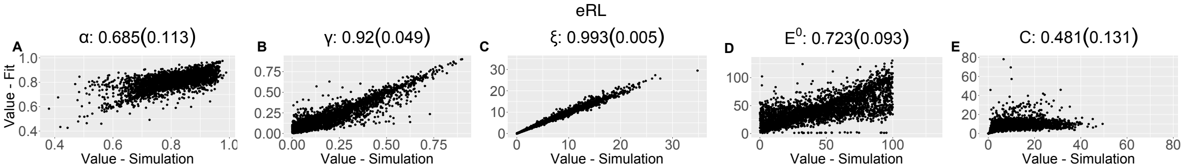


Figure S7: **Parameter recovery scatter plot for eRL model from** ≈ 100 **simulations.**

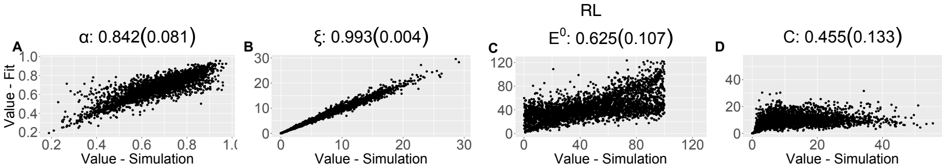


Figure S8: **Parameter recovery scatter plot for RL model from** ≈ 100 **simulations.**

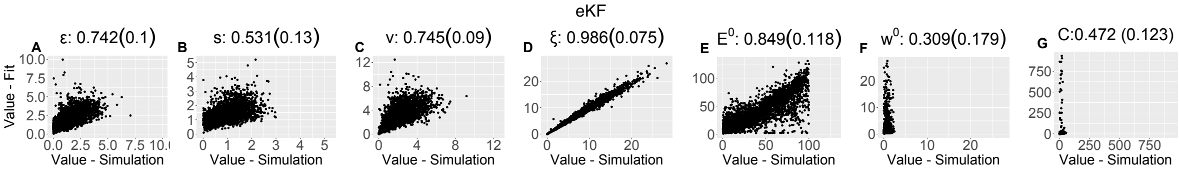


Figure S9: **Parameter recovery scatter plot for eKF model from** ≈ 100 **simulations.**

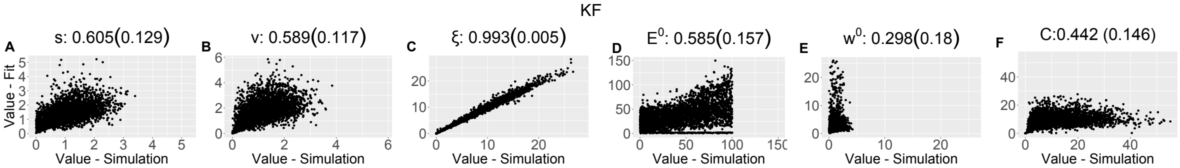


Figure S10: **Parameter recovery scatter plot for KF model from** ≈ 100 **simulations.**

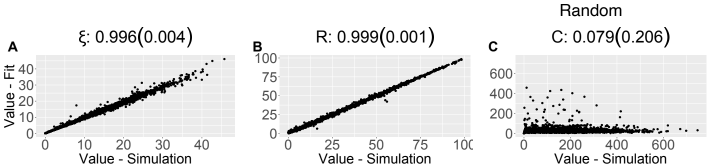


Figure S11: **Parameter recovery scatter plot for Random model from** ≈ 100 **simulations.**

### 5 Model recovery

We also ran model recovery analysis as described in the Methods. We report the confusion matrix of our analysis based on approximately 100 simulations (per model pair) in Table S4.

Table S4: Confusion matrix from the model recovery analysis based on ≈ 100 simulations. The y-axis indicates which model simulated the dataset, while the x-axis indicates which model fit the data based on LOOIC.

| Simulated \ Fit | eRL | RL | eKF | KF | Random |
|---|---|---|---|---|---|
| eRL | 0.327 | 0.173 | 0.404 | 0.096 | 0.000 |
| RL | 0.223 | 0.234 | 0.223 | 0.319 | 0.000 |
| eKF | 0.382 | 0.067 | 0.427 | 0.124 | 0.000 |
| KF | 0.229 | 0.281 | 0.281 | 0.208 | 0.000 |
| Random | 0.292 | 0.000 | 0.358 | 0.000 | 0.349 |

### 6 Condition-wise model comparison

For each condition, we ran model comparison procedure as described in the Methods. The results are reported in Table S5. In each condition, the expectation weighted models provided significantly better fit than models without this element.

Table S5: Model comparison results for each condition.

| Condition | Model name | ELPD difference | SE difference | Sigma effect | LOOIC |
|---|---|---|---|---|---|
| Vol. high Stoch. high | eKF - expectation weighted | 0.000 | 0.000 | | 15748.389 |
| | eRL - expectation weighted | -9.560 | 5.071 | 1.885 | 15767.509 |
| | RL | -139.407 | 61.362 | 2.272 | 16027.202 |
| | KF | -161.444 | 77.335 | 2.088 | 16071.277 |
| | Random response | -730.600 | 77.009 | 9.487 | 17209.588 |
| Vol. high Stoch. low | eKF - expectation weighted | 0.000 | 0.000 | | 15682.115 |
| | eRL - expectation weighted | -17.439 | 5.896 | 2.958 | 15716.993 |
| | RL | -131.817 | 35.936 | 3.668 | 15945.749 |
| | KF | -133.464 | 37.171 | 3.591 | 15949.042 |
| | Random response | -824.346 | 79.148 | 10.415 | 17330.807 |
| Vol. low Stoch. high | eKF - expectation weighted | 0.000 | 0.000 | | 15990.114 |
| | eRL - expectation weighted | -12.027 | 7.029 | 1.711 | 16014.169 |
| | RL | -149.338 | 43.874 | 3.404 | 16288.789 |
| | KF | -159.738 | 46.485 | 3.436 | 16309.590 |
| | Random response | -831.096 | 84.549 | 9.830 | 17652.306 |
| Vol. low Stoch. low | eKF - expectation weighted | 0.000 | 0.000 | | 15904.936 |
| | eRL - expectation weighted | -11.068 | 4.309 | 2.569 | 15927.072 |
| | RL | -70.588 | 16.643 | 4.241 | 16046.111 |
| | KF | -74.031 | 20.972 | 3.530 | 16052.997 |
| | Random response | -901.792 | 107.244 | 8.409 | 17708.519 |

### 7 Model diagnostics

In Table S6-S9, we report Bulk and Tail Effective Sample Size (ESS) for each condition, for each model and parameter.

Table S6: Bulk and Tail Effective Sample Size(ESS) Values for Vol. High - Stoch. High

| Model | Param. | ESS (Bulk) | ESS (Tail) |
|---|---|---|---|
| eRL | $\alpha$ | 58.166 | 47.491 |
| | $C$ | 90.5 | 79.142 |
| | $E^0$ | 54.655 | 137.729 |
| | $\xi$ | 31.233 | 47.726 |
| | $\gamma$ | 39.509 | 49.335 |
| RL | $\alpha$ | 56.22 | 36.057 |
| | $C$ | 99.642 | 52.599 |
| | $E^0$ | 126.757 | 467.373 |
| | $\xi$ | 31.322 | 36.92 |
| eKF | $C$ | 89.281 | 83.274 |
| | $E^0$ | 37.723 | 103.977 |
| | $\epsilon$ | 94.203 | 429.332 |
| | $v$ | 53.099 | 41.511 |
| | $s$ | 1665.566 | 4593.161 |
| | $w^0$ | 616458.467 | 467603.626 |
| | $\xi$ | 31.322 | 47.1 |
| KF | $C$ | 101.584 | 55.345 |
| | $E^0$ | 122.76 | 512.134 |
| | $v$ | 114.644 | 54.015 |
| | $s$ | 438.028 | 730.579 |
| | $w^0$ | 904.643 | 6759.804 |
| | $\xi$ | 31.457 | 36.763 |
| Random | $R$ | 27.939 | 33.982 |
| | $C$ | 397.862 | 259.967 |
| | $\xi$ | 32.334 | 41.271 |

Table S7: Bulk and Tail Effective Sample Size (ESS) Values for Vol. High - Stoch. Low

| Model | Param. | ESS (Bulk) | ESS (Tail) |
|---|---|---|---|
| eRL | $\alpha$ | 86.32 | 60.849 |
| | $C$ | 235.396 | 373.736 |
| | $E^0$ | 43.489 | 109.903 |
| | $\xi$ | 30.471 | 36.664 |
| | $\gamma$ | 42.125 | 55.178 |
| RL | $\alpha$ | 49.221 | 40.877 |
| | $C$ | 328.761 | 455.542 |
| | $E^0$ | 63.341 | 111.689 |
| | $\xi$ | 30.304 | 38.063 |
| eKF | $C$ | 227.813 | 363.944 |
| | $E^0$ | 33.393 | 104.395 |
| | $\epsilon$ | 376.691 | 1218.299 |
| | $v$ | 45.861 | 37.486 |
| | $s$ | 99526.69 | 148393.383 |
| | $w^0$ | 567627.288 | 634817.458 |
| | $\xi$ | 30.438 | 36.66 |
| KF | $C$ | 328.005 | 448.632 |
| | $E^0$ | 57.467 | 124.471 |
| | $v$ | 293.426 | 480.255 |
| | $s$ | 164.454 | 598.211 |
| | $w^0$ | 412979.973 | 354163.251 |
| | $\xi$ | 30.16 | 38.105 |
| Random | $R$ | 28.397 | 32.922 |
| | $C$ | 1794.614 | 1170.459 |
| | $\xi$ | 30.204 | 34.896 |

Table S8: Bulk and Tail Effective Sample Size(ESS) Values for Vol. Low - Stoch. High

| **Model** | **Param.** | **ESS (Bulk)** | **ESS (Tail)** |
|---|---|---|---|
| eRL | $\alpha$ | 43.312 | 40.66 |
| | $C$ | 248.885 | 434.44 |
| | $E^0$ | 49.006 | 85.409 |
| | $\xi$ | 29.68 | 34.909 |
| | $\gamma$ | 45.37 | 52.755 |
| RL | $\alpha$ | 39.911 | 35.351 |
| | $C$ | 433.949 | 435.575 |
| | $E^0$ | 181.442 | 618.317 |
| | $\xi$ | 29.527 | 36.192 |
| eKF | $C$ | 248.848 | 418.003 |
| | $E^0$ | 35.363 | 51.728 |
| | $\epsilon$ | 1272.838 | 2427.211 |
| | $v$ | 41.144 | 40.915 |
| | $s$ | 2399.657 | 6854.212 |
| | $w^0$ | 612283.163 | 531588.25 |
| | $\xi$ | 29.699 | 34.762 |
| KF | $C$ | 423.339 | 417.747 |
| | $E^0$ | 88.749 | 302.863 |
| | $v$ | 58.795 | 47.015 |
| | $s$ | 206.969 | 672.666 |
| | $w^0$ | 499152.469 | 573964.793 |
| | $\xi$ | 29.511 | 36.341 |
| Random | $R$ | 27.892 | 32.919 |
| | $C$ | 269.239 | 106.139 |
| | $\xi$ | 29.69 | 44.38 |

Table S9: Bulk and Tail Effective Sample Size(ESS) Values for Vol. Low - Stoch. Low

| **Model** | **Param.** | **ESS (Bulk)** | **ESS (Tail)** |
|---|---|---|---|
| eRL | $\alpha$ | 57.116 | 40.932 |
| | $C$ | 162.472 | 129.413 |
| | $E^0$ | 43.707 | 117.295 |
| | $\xi$ | 29.632 | 34.486 |
| | $\gamma$ | 65.497 | 151.548 |
| RL | $\alpha$ | 45.892 | 37.244 |
| | $C$ | 158.681 | 98.898 |
| | $E^0$ | 80.406 | 441.719 |
| | $\xi$ | 29.558 | 35.077 |
| eKF | $C$ | 149.16 | 126.209 |
| | $E^0$ | 38.88 | 73.732 |
| | $\epsilon$ | 653.635 | 1473.554 |
| | $v$ | 48.883 | 43.445 |
| | $s$ | 2263.547 | 9318.066 |
| | $w^0$ | 635517.969 | 313426.188 |
| | $\xi$ | 29.699 | 34.721 |
| KF | $C$ | 158.729 | 105.929 |
| | $E^0$ | 71.438 | 457.431 |
| | $v$ | 91.988 | 69.957 |
| | $s$ | 287.835 | 895.249 |
| | $w^0$ | 527620.655 | 587092.529 |
| | $\xi$ | 29.527 | 35.147 |
| Random | $R$ | 28.474 | 38.123 |
| | $C$ | 2426.581 | 1279.66 |
| | $\xi$ | 29.532 | 34.731 |

While some of the ESS values are below the recommended threshold of 100, indicating potential issues with parameter inference. This may be due to a low participant sample size, as well as small number of trials per condition, hinting limited statistical power. Given that the Rhat values are all around 1, and that there are no divergent transitions, as well as a fairly good parameter recovery, we see this as a minor issue.

Lastly, in Table S10 we model diagnostics for each condition, such as the Estimated Bayesian Fraction of Missing Information (E-BFMI), number of divergent transition, and E-BFMI values per chain.

Table S10: Model diagnostics for each condition - Estimated Bayesian Fraction of Missing Information (E-BFMI), number of divergent transition E-BFMI values per chain.

| **Condition** | **Model** | **# chains low E-BFMI** | **# div. transitions** | **E-BFMI Values** |
|---|---|---|---|---|
| HVHS | eRL | 0 | 0 | 0.696 0.713 0.695 0.691 |
| | RL | 0 | 0 | 0.76 0.748 0.771 0.806 |
| | eKF | 0 | 0 | 0.755 0.767 0.771 0.759 |
| | KF | 0 | 0 | 0.633 0.596 0.547 0.563 |
| | Random | 0 | 0 | 0.842 0.851 0.843 0.835 |
| HVLS | eRL | 0 | 0 | 0.689 0.76 0.69 0.689 |
| | RL | 0 | 0 | 0.624 0.688 0.688 0.685 |
| | eKF | 0 | 0 | 0.741 0.764 0.753 0.779 |
| | KF | 0 | 0 | 0.654 0.734 0.689 0.674 |
| | Random | 0 | 0 | 0.883 0.779 0.836 0.833 |
| LVHS | eRL | 0 | 0 | 0.73 0.732 0.728 0.702 |
| | RL | 0 | 0 | 0.719 0.714 0.742 0.7 |
| | eKF | 0 | 0 | 0.753 0.755 0.792 0.766 |
| | KF | 0 | 0 | 0.75 0.768 0.729 0.754 |
| | Random | 0 | 0 | 0.864 0.849 0.883 0.845 |
| LVLS | eRL | 0 | 0 | 0.764 0.762 0.75 0.764 |
| | RL | 0 | 0 | 0.714 0.764 0.719 0.697 |
| | eKF | 0 | 0 | 0.783 0.751 0.772 0.77 |
| | KF | 0 | 0 | 0.705 0.695 0.702 0.726 |
| | Random | 0 | 0 | 0.835 0.829 0.852 0.847 |

### 8 Modelling results

#### 8.1 Group-level differences between each condition

We plotted the estimate posterior distributions for each parameter of the model (including the across-trial average and the final learning rate and perceptual weighting term) in Figure S12. We found no group-level differences between conditions for any of the posterior distribution of the parameters in the winning eKF model.

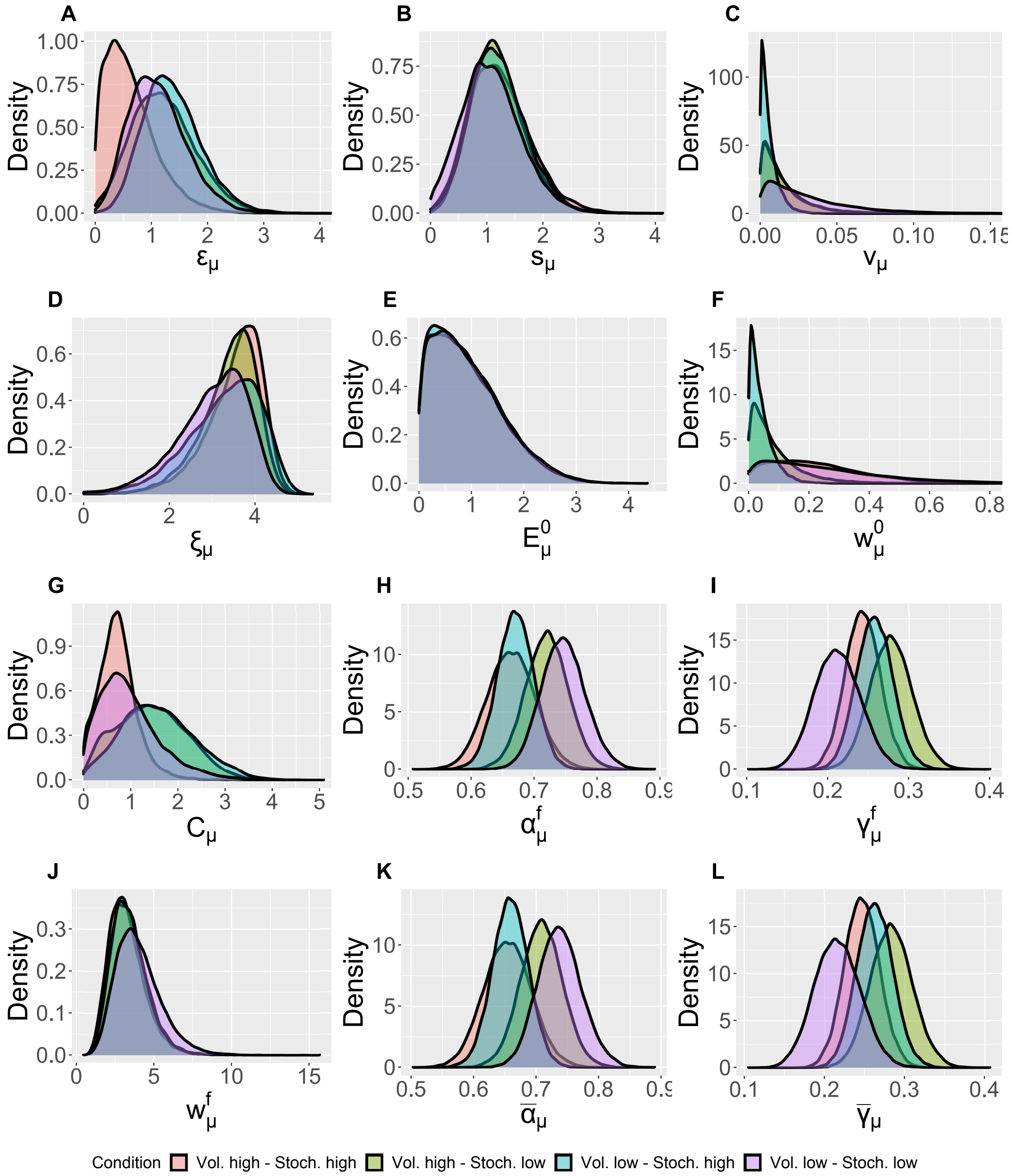


Figure S12: Group-level distributions for parameters for each condition for the eKF model.

#### 8.2 Individual-level differences between conditions

We estimated the individual-level parameters for each condition, we include their violin plots in Figure S13.

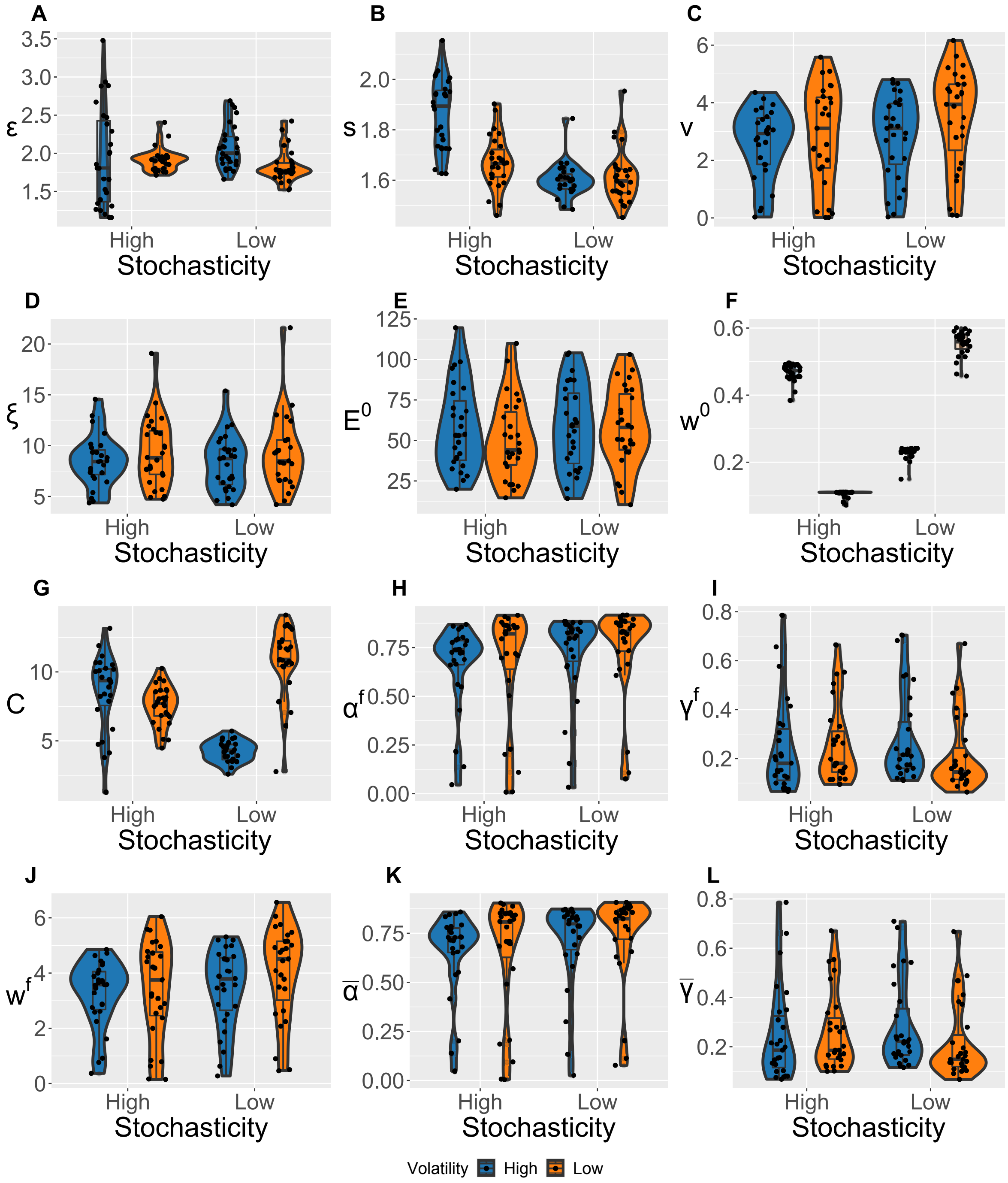


Figure S13: Violin plots (and box-plots) of individual-level parameters for each condition in the winning eKF model.
